# Mutational landscape of pure ductal carcinoma in situ and associations with disease prognosis and response to radiotherapy

**DOI:** 10.1101/2025.03.01.25323122

**Authors:** Noor Rizvi, Eliseos J. Mucaki, Emily L. Salmini, Monica Zhang, Sabina Trebinjac, Ezra Hahn, Lawrence Paszat, Sharon Nofech-Mozes, Michael T. Hallett, Eileen Rakovitch, Vanessa Dumeaux

**Author notes:** Corresponding author: Vanessa Dumeaux.

## Abstract

**Background:** Managing Ductal Carcinoma in Situ (DCIS) remains challenging due to the lack of reliable biomarkers to predict radiotherapy (RT) response, leading to both overtreatment of indolent disease and undertreatment of aggressive cases.

**Results:** Through whole-exome sequencing of 147 DCIS cases, we characterized the genomic landscape of pure DCIS and identified genetic alterations associated with the risk of recurrence, either in-situ or invasive. DCIS lesions harbored frequent mutations in established cancer drivers (*PIK3CA*, *TP53*) and genes regulating tissue architecture, which likely enhanced pre-invasive cell fitness but lacked prognostic value. A subset of younger patients exhibited distinct mutational processes characterized by high mutational burden, though these were not linked to recurrence risk. Across the cohort, five mostly mutually exclusive genes (*SH2B2*, *PDZD8*, *MYO7A*, *MUCL3*, *DNASE2B*), involved in cell adhesion, membrane organization, and DNA degradation, were significantly associated with 10-year risk of local recurrence. In RT-treated patients, we identified 27 additional mutated genes uniquely associated with recurrence, along with *SH2B2* and *MUCL3*. Most of these genes were involved in cytoskeletal regulation, cell adhesion, and cell-environment interactions. Mutations in metabolic regulators (*MGAM2* and *AADACL3*) and *REV1*, which mediates DNA damage tolerance, may impair cellular responses to RT-induced stress. Notably, we identified distinct genes prognostic for in-situ versus invasive recurrence: nineteen genes predominantly involved in tissue structural maintenance in in-situ relapse, and thirteen genes primarily affecting cell-cycle and genome-stability pathways in invasive progression. Copy number analyses revealed that pure DCIS exhibits molecular subtype-specific patterns characteristic of invasive disease, with novel alterations associated with recurrence, including three non-adjacent gains and five losses in regions harboring oncogenes, tumor suppressor genes, and genes regulating structural integrity, cell-cell adhesion and interactions.

**Conclusions:** While *TP53*, *PIK3CA*, and recurrent copy number alterations represent early events in tumorigenesis, they lack prognostic value in pure DCIS, underscoring the need for alternative biomarkers. Our findings identify key genetic alterations associated with local recurrence and RT resistance. We further uncovered distinct molecular programs underlying in-situ versus invasive recurrence, with mutations affecting tissue structural maintenance in in-situ relapse and cell-cycle/genome-stability pathways in invasive progression.

## Introduction

Ductal Carcinoma in Situ (DCIS) is a non-invasive, non-obligate precursor of invasive breast cancer characterized by clonal proliferation of neoplastic cells confined within the breast ducts [1]. DCIS is primarily detected through screening and diagnostic mammograms due to its characteristic presentation with microcalcifications. The widespread implementation of mammographic screening programs has led to a substantial increase in DCIS detection, with incidence rates rising from four to eleven cases per 100,000 women between 1993 and 2007 [2,3]. This trend is expected to continue as screening programs expand to include younger women [4].

There are currently no definitive markers to predict which cases will progress to invasive, life-threatening lesions [5,6]. Consequently, treatment, usually involving breast-conserving surgery (BCS) followed by breast radiation therapy (RT), is recommended for all women diagnosed with DCIS [5]. However, many DCIS would not transit to invasive life-threatening disease even if left untreated [7]. The absence of known markers predictive of a patient benefit to RT results in both over-treatment for indolent lesions and under-treatment for some aggressive DCIS likely to evolve to an invasive state. Markers that identify such cases could enable tailored treatments, such as full mastectomy or adjuvant systemic therapies for patients at risk of in situ or invasive local recurrence (LR) despite receiving RT [8], or more frequent watch-and-wait strategies for those with indolent lesions.

Cancer emerges from the accumulation of genetic aberrations in neoplastic cells and genomic instability [9]. Mutations can disrupt normal cellular processes, leading to uncontrolled cell growth, impaired DNA repair, and other aberrant cellular behaviours that may contribute to cancer development [9]. Previous studies have revealed important insights into DCIS progression by highlighting specific mutations and chromosomal alterations that may influence the progression towards invasive disease [10–17]. Many of these studies, however, focus on synchronous DCIS – concurrent presentation of DCIS and invasive ductal carcinoma (IDC). These studies explore the overlap between DCIS and IDC, and recognize the potential differences between them. However, these findings might be reflective of a timepoint beyond the evolutionary bottleneck, and the full repertoire of mediators of the transition from DCIS to IDC or associated with a future risk of LR cannot be established from these studies [10].

*TP53* and *PIK3CA* [11–17] are among the most frequently observed mutations in these studies [11–17] and are often characterized as key drivers of tumorigenesis, promoting growth and spread of cancer cells. However, this may not necessarily be the case, as mutant field clonalization could equip pre-malignant cells with additional (epi-) genetic mutations that confer fitness advantages, allowing them to expand into ductal regions without directly driving invasive behavior [10]. This suggests that driver mutations, alongside frequently mutated genes, may serve as these early fitness enhancers rather than drivers of progression. Several copy number aberrations (CNAs) have also been identified by these studies [10–15,17–20] including frequent gains at 1q, 8q, 11q, and 17q, and losses at 16q, however the specific associations of these CNAs to DCIS prognosis remain unclear.

To address these critical gaps, we conducted comprehensive exome sequencing analysis of 147 pure DCIS cases, including patients treated with and without RT, to investigate markers of local recurrence within 10 years of diagnosis. Our study characterizes the mutational landscape in pure DCIS and identifies novel genomic alterations associated with tumor grade, molecular subtypes, and patient age. Most importantly, we discovered specific variants and CNAs predictive of local recurrence risk, including genetic markers associated with RT response. These findings provide insights into the genomic determinants of DCIS prognosis and treatment response, establishing a foundation for improved risk stratification and personalized treatment strategies for DCIS patients.

## Results

### A unique cohort of pure DCIS patients

We assembled a cohort of 147 pure DCIS patients treated with BCS, with or without subsequent RT, incorporating comprehensive genomic profiling through whole-exome DNA analysis of primary DCIS tumors and matched normal tissues (**Table 1**). The study design was balanced to include at least a third of patients who experienced an ipsilateral invasive or in-situ local recurrence within a 10-year follow-up period and about half received radiotherapy as part of their standard-of-care (**Table 1**). In clinical settings, RT is omitted in some patients with low-risk features of DCIS or due to patient preference. The median time to recurrence was 4.2 years for invasive disease and 2.1 years for in-situ disease. Most tumors were of intermediate to high grade spanning all five molecular subtypes, with normal-like and luminal A subtypes more frequently observed in patients without local recurrence within 10 years (**Table 1**). A minority of tumors exhibited multifocality (24.5%) and positive margins (6.8%). Clinical characteristics were comparable between RT-treated and untreated patients across all variables except age, with women aged 60 years or older less likely to receive RT (**Supplementary Table 1**).

**Table 1.**
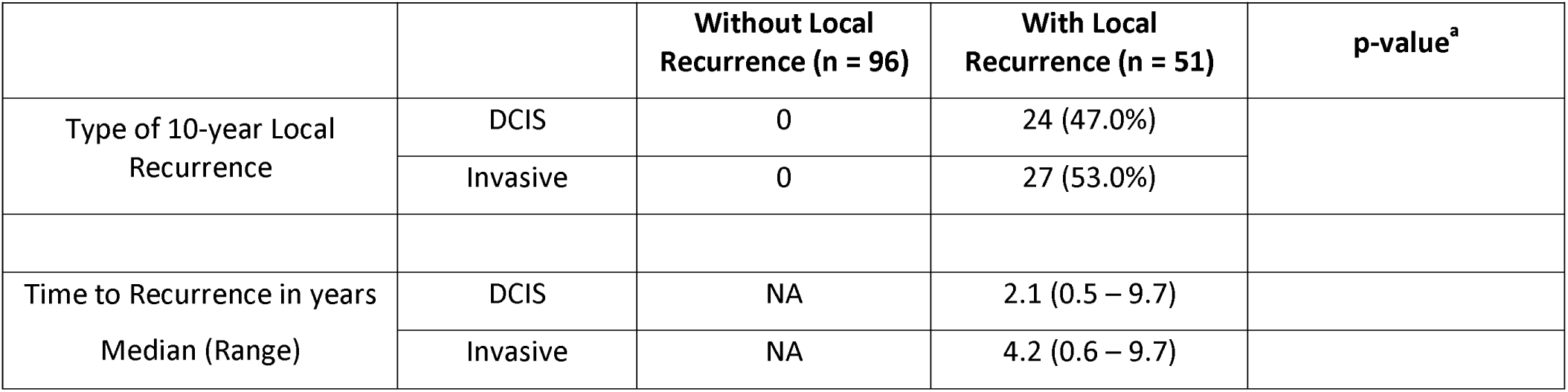
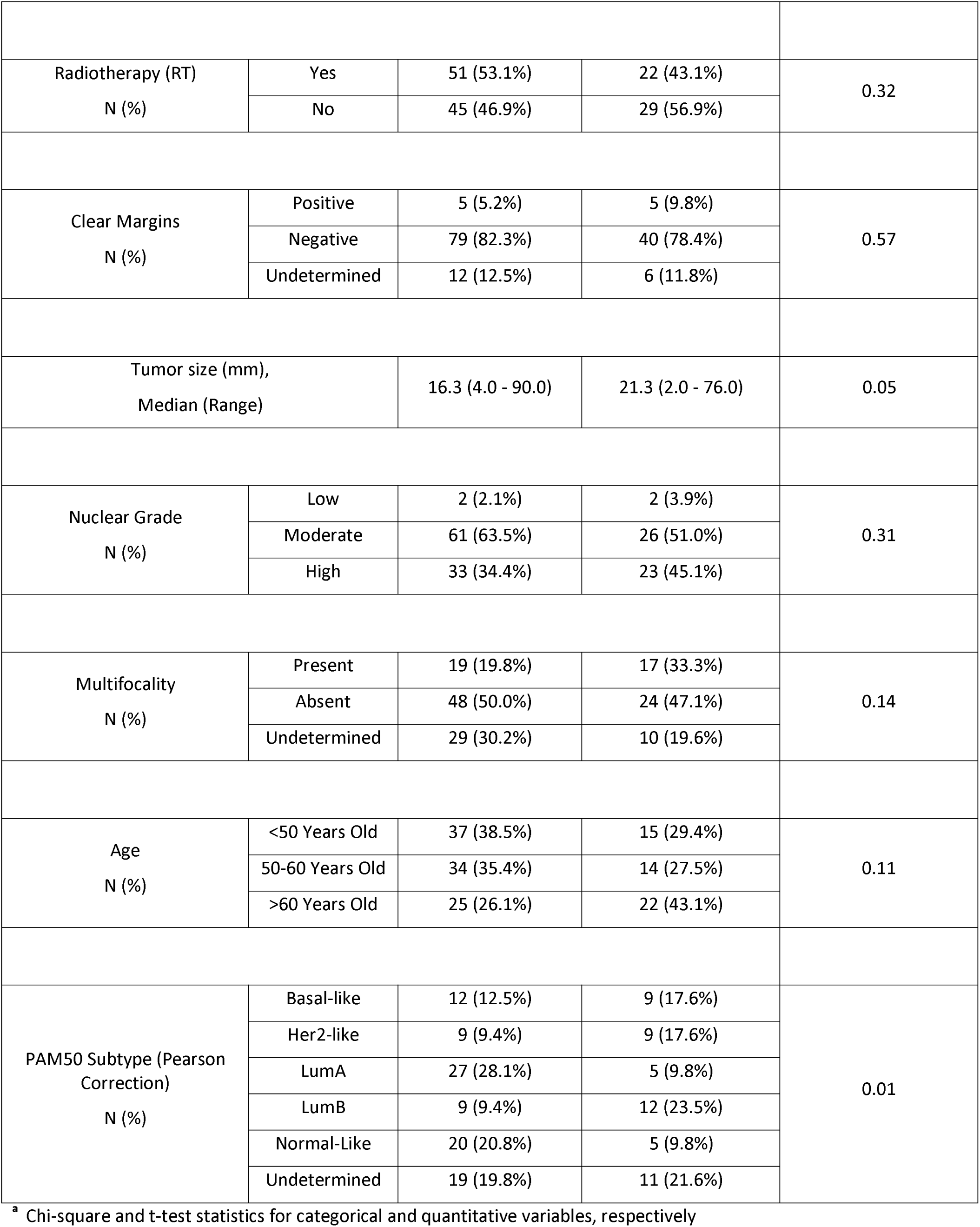
Patient and tumor clinical attributes.

### Mutational landscape of DCIS reveals distinct processes driving high mutational burden in early-onset cases

The mutational load of pure DCIS lesions varied considerably, ranging from. 3 to 3,482 non-synonymous variants per sample (median 75) The majority (74.2%) were missense mutations, constituting approximately 52K unique variants identified in ∼14K genes. Most of these genes were not frequently mutated, with 2,030 genes (14.5%) harboring non-synonymous aberrations in at least 5 patients.

As expected, most mutations are of C>T type (barplot in **Fig. 1**), a common mutational pattern attributed to the spontaneous deamination of 5-methylcytosine, a process frequently observed in many cancer genomes [21,22]. Spontaneous deamination can be exacerbated in FFPE samples due to DNA damage [23]. FFPE-related mutational artefacts are known to resemble certain COSMIC signatures [24], such as SBS30 and SBS1 [25]. During library preparation, formalin-induced DNA lesions are chemically repaired with unrepaired profiles resembling SBS30 and repaired profiles resembling SBS1 [25]. While SBS30 was not identified in our dataset, SBS1 was detected in a large number of samples (**Fig. 1**). SBS1, commonly found in tumor genomes, is difficult to distinguish from repaired FFPE-related artifacts due to their high similarity [22,25]. However, previous research has demonstrated that mutational profiles from repaired FFPE samples closely match true tumor mutational profiles [25], and we expect that our careful processing of mutation calling-pipeline minimizes the impact of these artefacts [23].

**Fig. 1:**
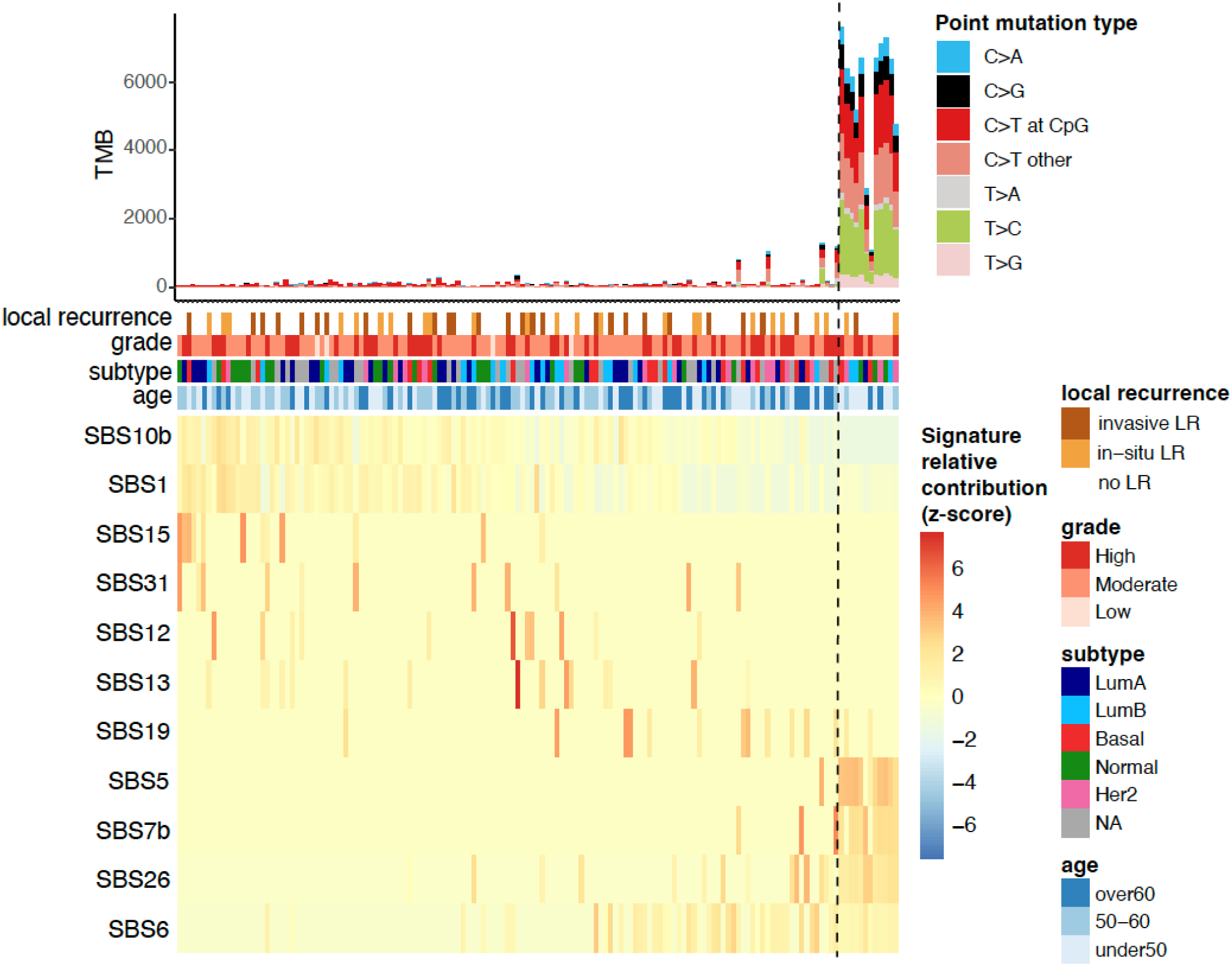
**Mutational Signatures in pure DCIS samples.** The top panel shows tumor mutational burden (TMB) with distribution of point mutation types. Clinical annotations display 10-year local recurrence outcome, tumor grade, molecular subtype, and patient age. The bottom heatmap depicts the row-scaled relative contribution of single base substitution (SBS) signatures detected in at least 10 samples (z-score). Samples (columns) and signatures (rows) are ordered based on a rank-sum statistic that maximizes the coherence of contribution patterns. The method first splits signatures into two groups using medoid clustering. For each signature, relative contributions are ranked within each group, and the final sample ordering is determined by the average rank-sum across all signatures.

Interestingly, a subset of DCIS samples exhibit a significantly higher tumor mutational burden (TMB), characterized by an increase in frequency of C>G, T>C, and C>A mutations, and enrichment of specific SBS signatures including SBS26, SBS7b, and SBS5 (**Fig. 1**, 12 samples on the right). The causes underlying these mutational signatures remain incompletely understood. However, SBS26 has been linked to impaired mismatch repair and microsatellite instability. These cases were not more likely to have a recurrence, of a higher grade or of a specific molecular subtype but were predominantly found in younger patients (8 out of 12 under 50 years; Chi-square test p-value < 0.05).

These findings highlight distinct mutational processes in some early-onset DCIS cases, though these molecular features were not associated with prognosis.

### Pure DCIS is associated with high frequency of mutations in genes involved cell adhesion, polarity, tissue structure and function

Analysis of pure DCIS revealed distinct patterns of recurrent mutations across multiple genes (**Fig. 2**). *PIK3CA* was the most frequently mutated gene (15% of cases), followed by *FSIP2* and *KIR3DL3* (14%). We also identified functional gene groups among the most frequently mutated genes including several motor genes converting chemical energy to mechanical force (*DNAH12, DNHD1*, and *MYOB15;* 12% each), collagen genes (*COL18A1* and *COL4A3*; 12% and 10%, respectively), and mucin genes (*MUC3A*, *MUC4*, *MUC22*, and *MUC5AC*) also showed alterations (10-12% of cases each). While mucin genes are typically large and can accumulate mutations by chance, these specific mucin genes were not identified as FLAGS (FrequentLy mutAted GeneS) and therefore not excluded in our analysis (See Methods). Given our stringent rules for mutation calling, these findings suggest that mutations in mucin and other genes controlling epithelial-components including cellular morphology, epithelial function and adhesion are central to the physiopathology of pure DCIS.

**Fig. 2:**
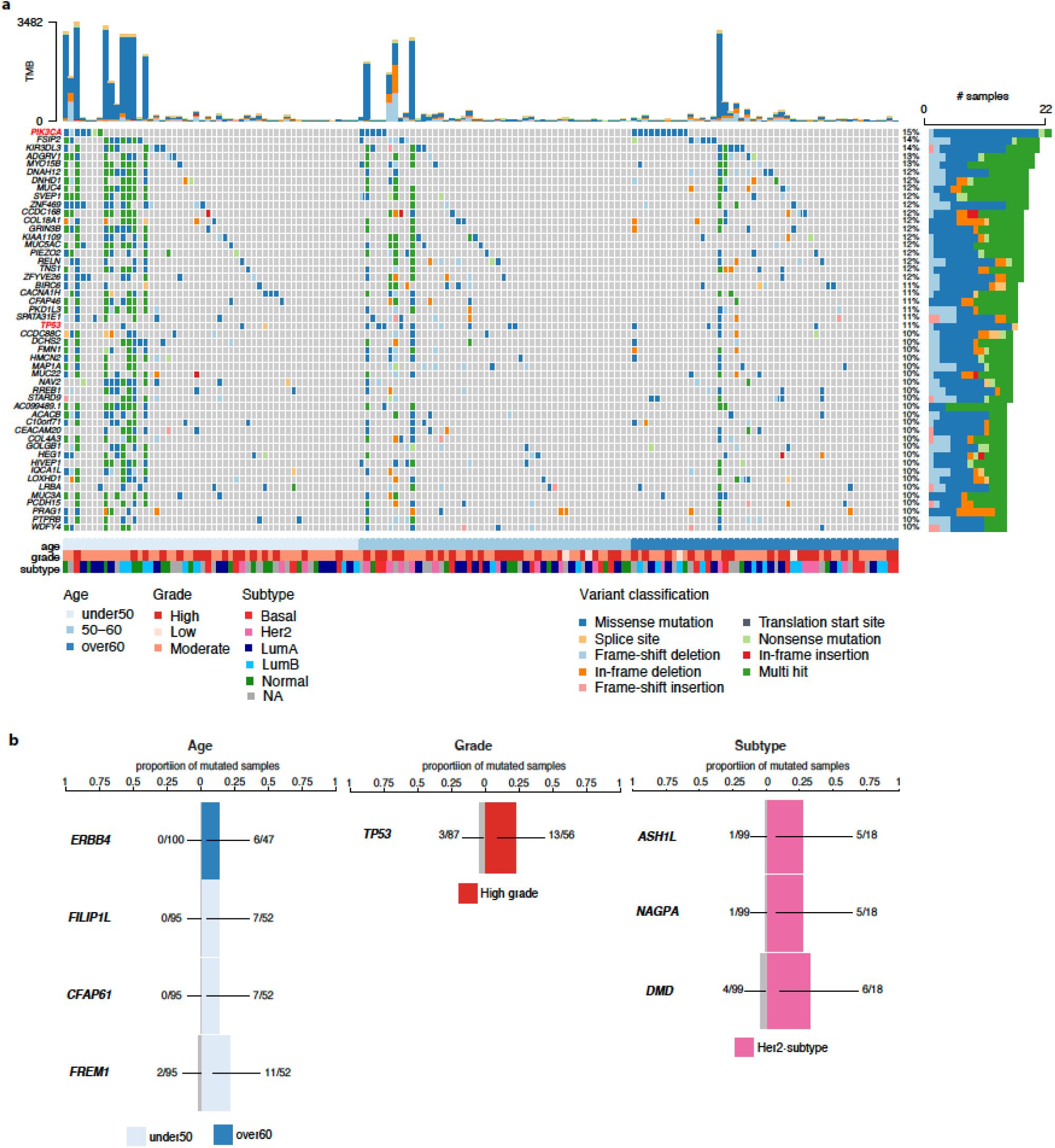
The most frequently mutated genes and association with clinico-pathological variables in pure DCIS patients. **a** The top 50 most frequent non-synonymous small variants identified in pure DCIS lesions. Samples are in columns and variants are color-coded based on their classification. The TMB for each lesion is displayed at the top of the heatmap. Samples are organized by age at diagnosis, with additional clinico-pathological features—grade and subtype—depicted at the bottom. **b** Mutated genes significantly associated with specific clinico-pathological variables (False discovery rate [FDR] < 0.001). The barplots show the proportion of mutated samples within specific categories: age group (left), high grade (middle), and Her2-subtype (right).

We also identified a few mutated genes enriched in distinct clinico-pathological groups including patients with early-onset DCIS (*FILP1L*, *CFAP61*, *FREM1*) or later-onset (*ERBB4*), high-grade lesions (*TP53*) and Her2-enriched subtype (*ASH1L*, *NAGPA*, *DMD*) (Figure 2B).

Collectively, these findings highlight that pure DCIS harbors frequent mutations in genes governing tissue architecture and cell-cell interactions, suggesting these alterations may be fundamental to DCIS development.

Proportions for each category are compared to the proportions of mutated samples in the other respective groups (shown in grey).

### Established cancer driver genes are present in pure DCIS but lack prognostic capacity

To identify potential cancer driver genes, we analyzed the ratio of non-synonymous to synonymous mutations (dN/dS) across all genes, which can indicate positive selection of mutations that provide growth advantages to cancer cells [26]. This analysis identified two significantly mutated driver genes in pure *DCIS*: *PIK3CA* and *TP53* (FDR < 0.05). Both genes were also among the most frequently mutated genes (Fig. 2a, red-labelled genes). While PIK3CA mutations were not enriched in specific molecular subtypes, TP53 mutations were significantly more frequent in basal-like and HER2-enriched subtypes compared to other subtypes (19% in basal-like [4/21], 33% in HER2-enriched [6/18] vs 5% in other subtypes [4/78], Chi-square test p < 0.005).

Out of 34 samples carrying a mutation in at least one of these driver genes, 15 samples harbored at least one potentially actionable alteration as indicated by OncoKB (**Supplementary Fig. 1**). The mutation spectrum in *PIK3CA* was dominated by the activating H1047R hotspot mutation (∼50%) in the kinase domain of exon 21, known to enhance PI3K-mitigated pathway signaling (**Supplementary Fig. 1a**) [27,28]. Similarly, *TP53* mutations clustered in known hotspots within the DNA-binding domain, which may result in the loss of tumor suppression by affecting its ability to bind to DNA (**Supplementary Fig. 1b**).

Despite their established roles in cancer progression, none of these driver mutations was associated with 10-year local recurrence risk (Firth’s penalized likelihood Cox regression p > 0.5), suggesting that additional factors influence DCIS prognosis.

### Identification of mutations associated with increased local recurrence risk independent of treatment

To identify potential prognostic markers, we analyzed mutated genes associated with the risk of invasive or in-situ local recurrence in the ipsilateral breast occurring between 6 months and 10 years after diagnosis. Our survival analysis identified five biomarkers each significantly associated with increased 10-year local recurrence risk (Firth’s penalized likelihood Cox regression p-value < 0.01 & permuted p-value < 0.05; **Fig. 3a-b**). These mutations, occurring in 4-7% of cases, were largely mutually exclusive and occurred across molecular subtypes and grades.

**Fig. 3:**
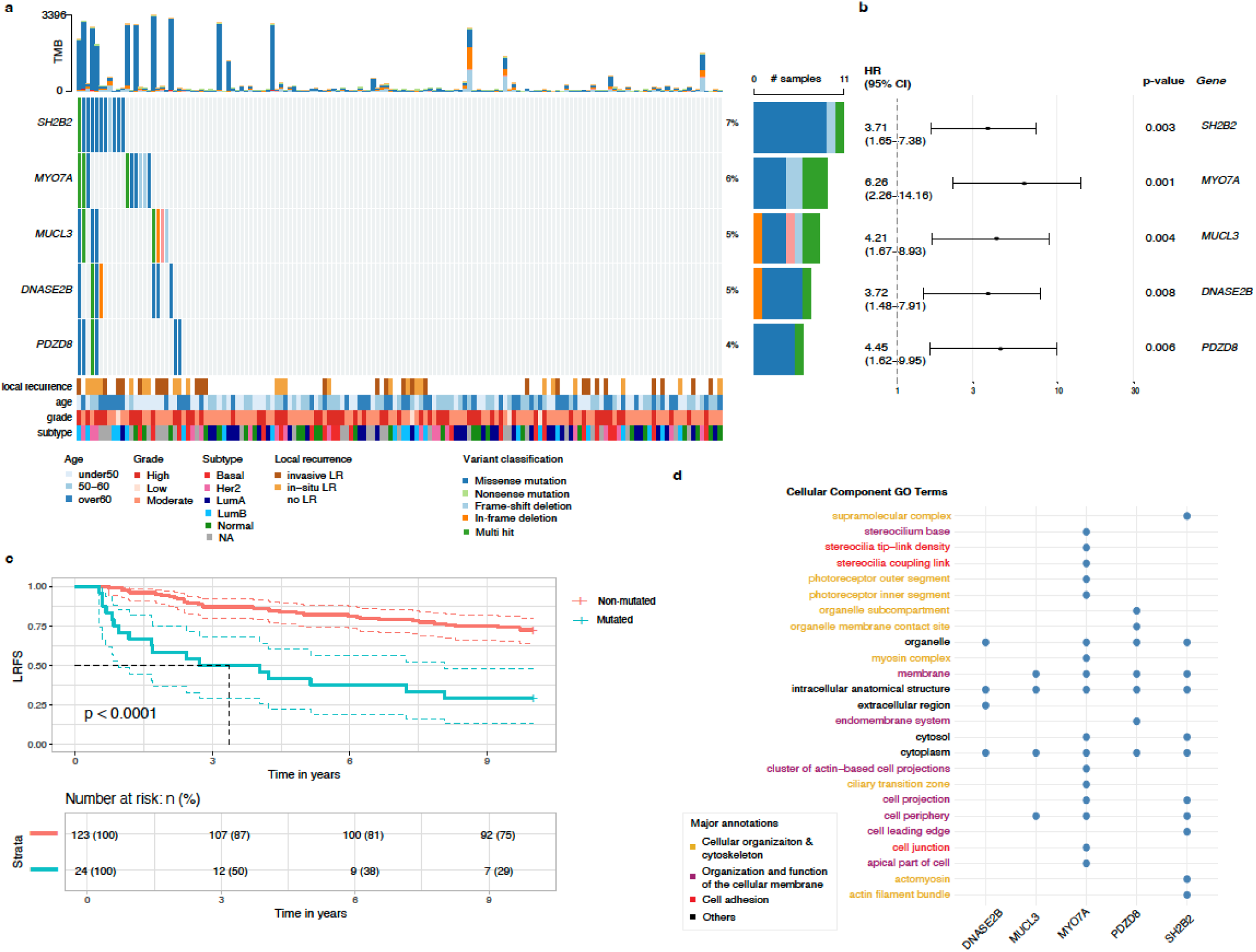
Gene variants associated with an increased 10-year local recurrence risk in DCIS. **a** Samples are in columns and variants are color-coded based on their classification. Local recurrence status and clinico-pathological characteristics of each lesion are depicted at the bottom. Local recurrence is defined as any recurrence, in situ or invasive, in the ipsilateral breast occurring between 6 months and 10 years after diagnosis. Right-hand side reports the proportion of each variant classification type. **b** Hazard ratio and confidence intervals for each significant mutated gene associated with increased 10-year local recurrence risk (Firth’s penalized likelihood Cox regression). **c** Kaplan-Meier analysis of local recurrence-free survival (LRFS) comparing patients with mutations in at least one of the 5 genes (red) versus those without mutations (blue). **d** Cellular component Gene Ontology (GO) terms annotations for each significant gene.

The presence of mutations in at least one of these genes was significantly associated with increased recurrence risk (**Fig. 3c**, log-rank p-value p<0.0001), with *MYO7A* and *PDZD8* showing the strongest associations (Firth’s penalized likelihood Cox regression HR>4.4, **Fig. 3b**). Gene Ontology analysis revealed that most genes are involved in multiple interrelated cellular processes including cell adhesion (*MYO7A*, *SH2B2*), organization and function of the cellular membrane (*PDZD8*, *MUCL3*, *MYO7A*), and cellular organization and cytoskeleton (*MYO7A*, *PDZD8*) (**Fig. 3d**). Additionally, DNASE2B, a member of the DNase II family of endonucleases, was identified among the significant genes. These findings underscore how cytoskeletal reorganization, changes in cell structure, and compromised cell adhesion might contribute to increased risk of recurrence within 10 years after a DCIS diagnosis.

### Mutations in genes governing cytoskeletal organization and membrane dynamics associated with radiotherapy resistance

To identify predictive biomarkers for RT response, we analyzed mutations associated with 10-year local recurrence in a cohort restricted to patients who received RT. This analysis revealed 29 genes significantly associated with an increased risk of recurrence (F<u>irth’s penalized</u> <u>likelihood Cox regression</u> p-value < 0.05 & permuted p-value < 0.05; **Fig. 4a**). Notably, these mutations often co-occurred, with at least two mutated genes present in approximately 27% of lesions (20/73 patients treated with RT).

**Fig. 4:**
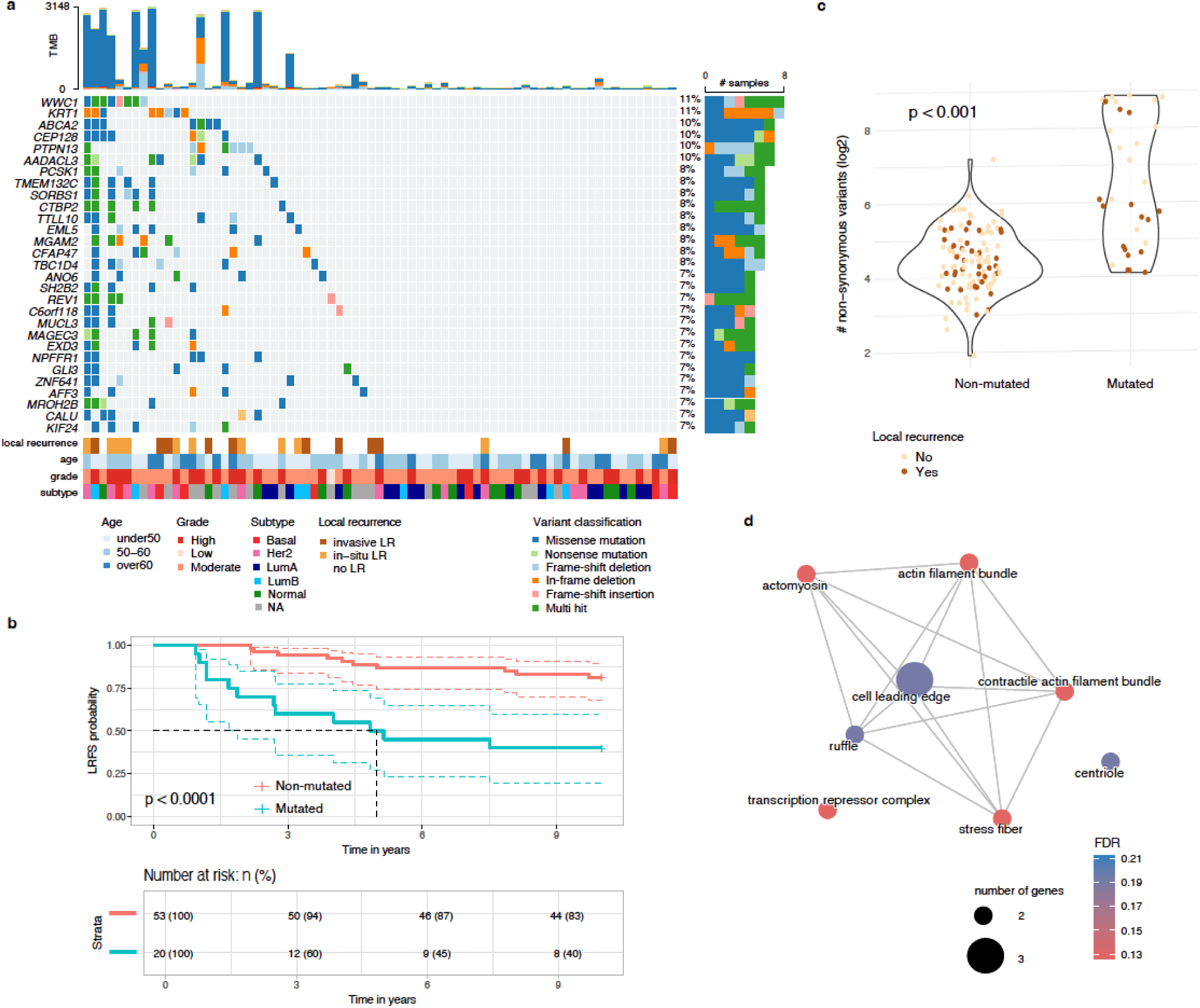
Mutations associated with radiotherapy resistance in DCIS. **a** An oncoplot showing the distribution of mutations in 29 genes significantly associated with local recurrence in RT-treated patients. Color-coding indicates mutation types; clinical annotations show age, grade, molecular subtype, and 10-year recurrence status. **b** Kaplan-Meier analysis of LRFS comparing patients with mutations in at least two of the 29 genes (red) versus those without mutations (blue). **c** Distribution of TMB, shown as number of non-synonymous variants, log2 scale) for lesions with at least one mutation in two genes associated with LRFS colored by recurrence status at 10 years. **d** GO cellular component enrichment network analysis of recurrence-associated genes in RT-treated DCIS. Network visualization shows enriched GO terms (FDR < 0.2) from genes linked to local recurrence following RT. Nodes represent individual GO terms with size proportional to gene count (>2), and edges indicate significant semantic similarity between terms. Node color intensity corresponds to enrichment significance.

In RT-treated patients, lesions harboring mutations in at least two of these genes exhibited a markedly increased risk of local recurrence (Firth’s penalized likelihood Cox regression HR = 4.8, 95% CI: 2.1–11.2, p = 0.0002), with most mutation-positive recurrences occurring within 5 years post-RT (**Fig. 4b**). Notably, while mutations in *SH2B2* and *MUCL3* were significantly associated with prognosis in both the overall cohort and the RT-treated subgroup, an RT-stratified analysis revealed that lesions harboring two mutations in the remaining 26 genes (which were altered in at least five lesions in the no-RT group) were predictive of prognosis only in RT-treated patients (p < 0.0001) and not in those who did not receive RT (p > 0.5) (**Supplementary Fig. 2**).

Given the frequent co-occurrence of these mutations, we assessed whether overall TMB might explain the increased recurrence risk. Although several high-TMB lesions were observed among patients with these mutations (**Fig. 4a,c**), TMB itself was not significantly associated with local recurrence risk (Firth’s penalized likelihood Cox regression p = 0.8). Indeed, many lesions with high TMB remained recurrence-free over 10 years, suggesting that specific mutations—not overall mutation load—drive LR after RT. We further examined potential confounding by clinico-pathological variables (i.e. age, grade, tumor size, multifocality and subtype) by adjusting our survival model for each factor. The predictive value of these mutations remained significant (Firth’s penalized likelihood Cox regression p-values < 0.005), supporting their role as an independent risk factor for recurrence following RT.

Functional analysis revealed a network of interrelated cellular functions involved in actin cytoskeleton regulation, cellular polarity, and membrane dynamics—processes essential for invasive behavior (**Fig. 4d**).

Alterations in *SH2B2* and *SORBS1*, which modulate stress fiber formation and actin remodeling, and in *PTPN13* and *WWC1*, which affect actomyosin tension at the cell leading edge, suggest disruptions in cytoskeletal dynamics. Additional genes associated with prognosis in RT-treated include *KRT1*, *MUCL3*, *TMEM132C*, *TTLL10*, *EML5*, *CEP128*, *CFAP47*, and *KIF24*, all of which contribute to maintaining cellular architecture, polarity, and adhesion (**Fig. 4a**). In addition, alterations in metabolic regulators—such as *MGAM2* and *AADACL3*— could influence the energetic and biosynthetic demands required in response to RT-induced stress, while *REV1* is implicated in DNA damage tolerance, a critical safeguard following genotoxic stress.

Collectively, these findings highlight the pivotal roles of cytoskeletal reorganization, membrane remodeling, metabolic regulation, and DNA repair pathways in the response to RT. The prognostic significance of these mutations in RT-treated patients suggests that these gene alterations may mediate resistance mechanisms triggered by RT-induced stress.

### Differential mutational associations with in-situ versus invasive local recurrence after radiotherapy

To investigate whether individual mutations are preferentially linked to either in-situ or invasive LR, we refitted the gene-based survival models using each LR type as the sole endpoint (the alternative event was censored) and stratified the analyses by RT. For the full cohort we had retained genes with p < 0.01 and permutation p < 0.05; for the smaller RT-treated group we used p < 0.05 and permutation p < 0.05. As in the main analysis, many significant associations were found in RT-treated patients, with some overlap with significant genes found in the whole cohort (**Fig. 5a**).

**Fig. 5:**
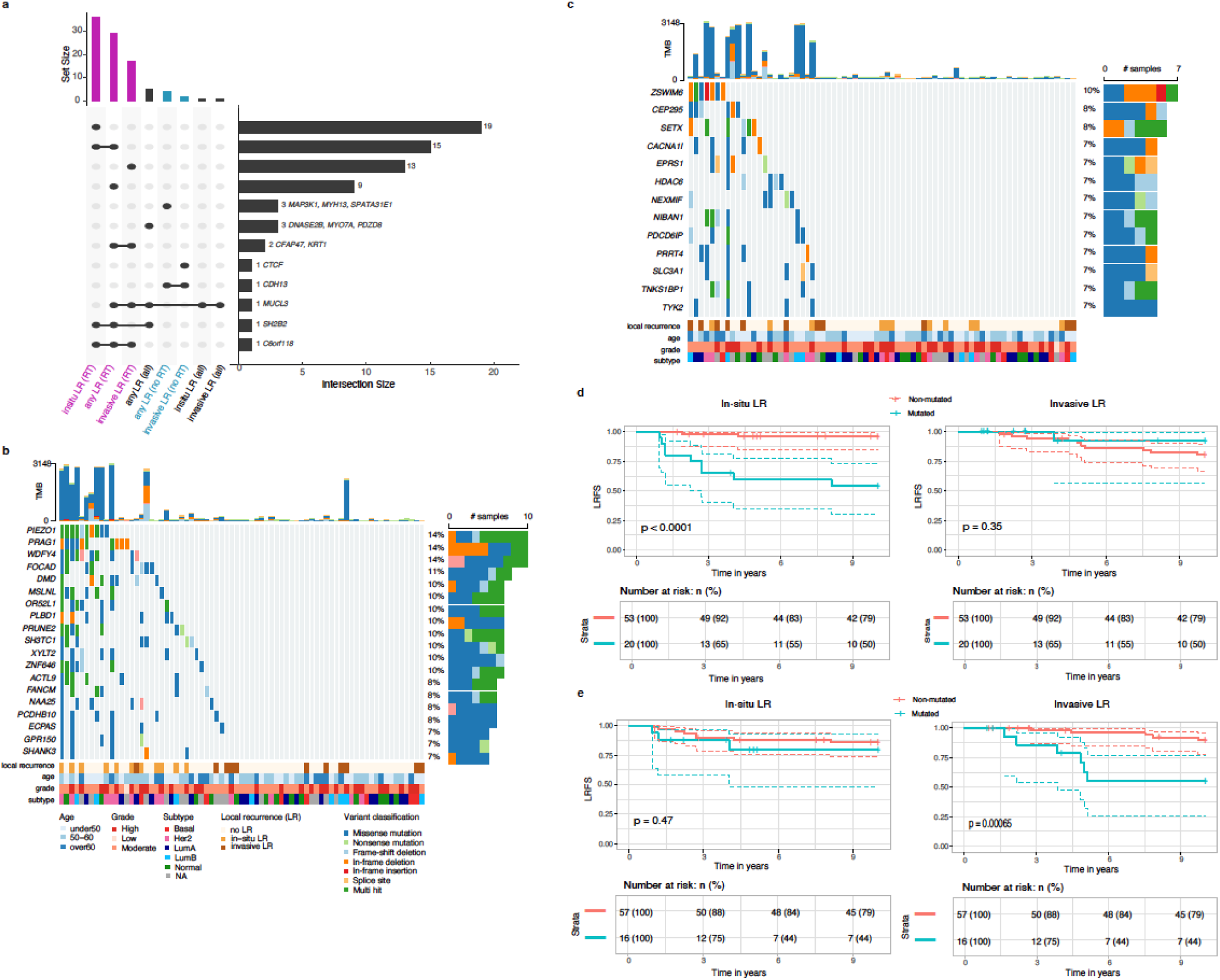
Differential mutational associations with in-situ versus invasive local recurrence after radiotherapy. **a** An UpSet plot showing the number of genes significantly associated with risk of invasive, in-situ or any local recurrence (LR) in the whole cohort (all) and in RT-treated (RT) or untreated patients (noRT), with intersections between gene lists indicated by connected dots below and corresponding size of the intersection depicted in the barplot on the right. **b** Oncoplot showing mutations in genes uniquely associated with in-situ LR (n=19) in RT-treated patients. Color-coding indicates mutation types; clinical annotations show age, grade, molecular subtype, and recurrence status. **c** Similar to b but showing mutations in genes uniquely associated with invasive LR (n=13) in RT-treated patients. **d** Kaplan-Meier analysis of in-situ (left) or invasive (right) LR-free survival comparing patients with mutations in at least two of the 19 genes uniquely associated with in-situ LR (red) versus those without mutations (blue). **e** Similar to d but comparing in-situ or invasive LR-free survival between patients with mutations in at least two of the 13 genes uniquely associated with invasive LR versus those without mutations.

Three genes—*MUCL3*, *SH2B2* and *C6orf118*—were associated with outcome in at least three out of six analyses (**Fig. 5a**). In RT-treated tumours, fifteen genes were associated with in-situ LR or with “any LR”, and two genes (*CFAP47*, *KRT1*) were associated with invasive or with “any LR” (**Fig. 5a**). Nineteen additional genes were uniquely associated with in-situ LR (**Fig. 5b**), whereas thirteen were unique to invasive LR (**Fig. 5c**) within the RT subgroup. The Kaplan-Meyer analyses demonstrate that patients whose lesions harbor mutations in at least two genes from either the in-situ-specific or invasive-specific gene sets show significantly worse LR-free survival only for the corresponding recurrence type (**Fig. 5d,e**).

Functional annotation indicates that mutations uniquely linked to in-situ LR occur mainly in genes that preserve epithelial architecture and mechanosensing: anchoring-junction components that couple cells to neighbouring cells or to the extracellular matrix (*DMD*, *FOCAD*), the stretch-activated channel PIEZO1, and scaffolds/adaptors localised to actin-rich membrane projections such as invadopodia (*SHANK3*, *PRAG1*). The set also includes WDFY4, a WD-repeat/FYVE-domain autophagy adaptor implicated in MHC-II antigen presentation.

Conversely, mutations associated with invasive LR after RT mapped to genes involved in late cytokinetic abscission (*PDCD6IP*), centriole-to-centrosome maturation (*CEP295*), DNA-damage sensing and repair (*SETX*, *TNKS1BP1*), calcium-regulated motility (*CACNA1I*) and cytokine-dependent inflammatory signalling (*TYK2*).

Because stratifying simultaneously by treatment and LR type reduces sample size and event numbers, we cannot exclude the possibility that some endpoint-specific associations—or the absence of others—reflect differences in statistical power. Nevertheless, these findings suggest that distinct biological programs—structural maintenance versus cell-cycle, genome-stability and inflammatory pathways—underlie in-situ and invasive patterns of recurrence after RT.

Copy number alterations display molecular subtype-specific patterns with select genomic regions linked to local recurrence

Analysis of copy number alterations (CNAs) revealed recurrent chromosomal changes similar to those reported in invasive breast cancer (**Fig. 6a**). Significant gains were identified on chromosomal arms 1q, 8q, 16p, 17q, 20p, and 20q, while losses predominantly occurred on 8p, 9p, 11q, 13q, 14q, 16q, and 17p (binomial test, FDR < 0.05).

**Fig. 6:**
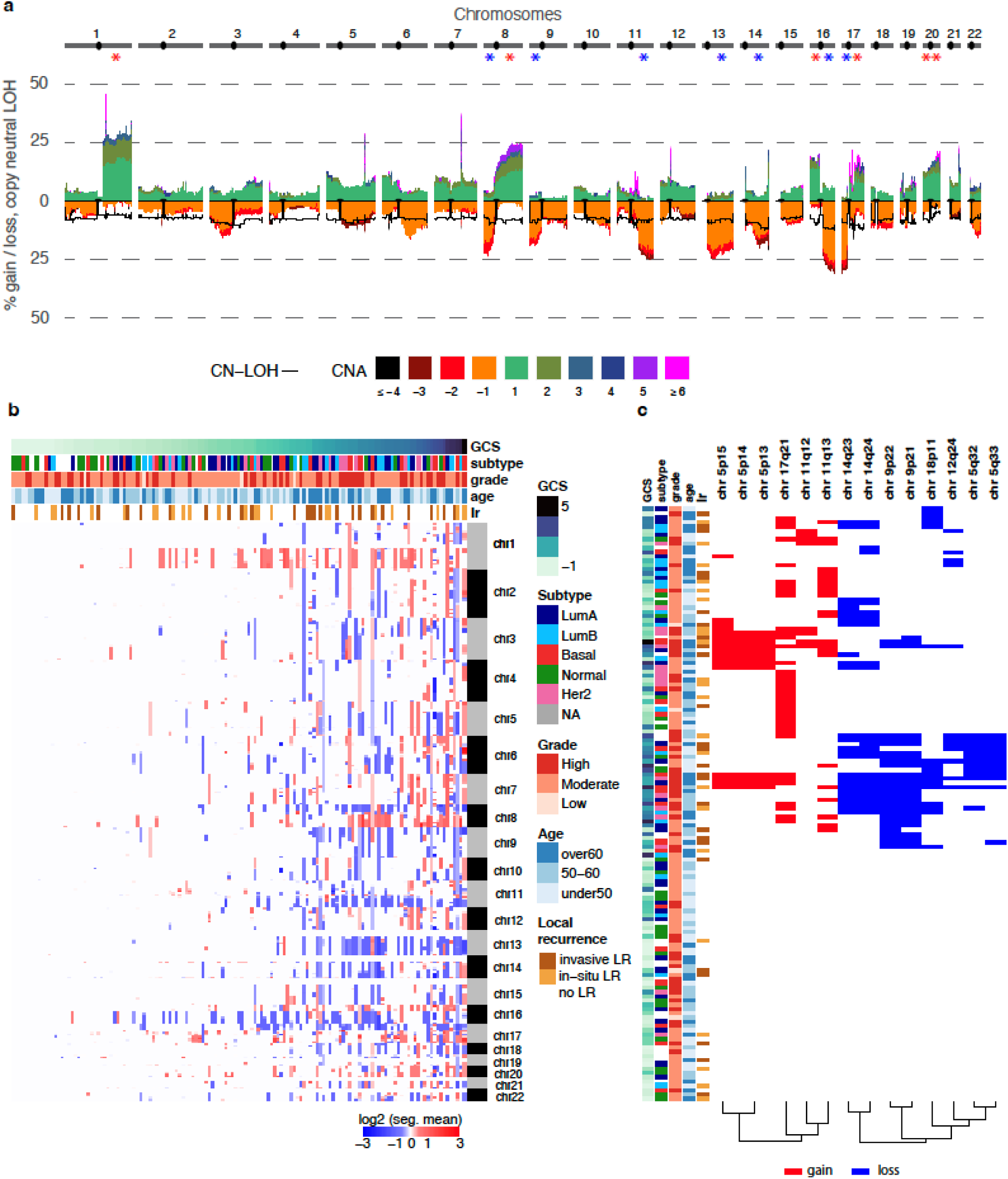
Frequent copy number alterations in pure DCIS and associations with 10-year LR risk. **a** Genome-wide frequency of absolute copy number gains and losses across chromosomes 1-22 (1Mb window). The estimated ploidy for each sample is subtracted from the copy number values of each segment which means that a copy number of 0 is no copy number change. The y-axis shows the percentage of samples with each alteration type. Asterisks indicate statistical significance for chromosomal arm alterations **b** Heatmap showing copy number profiles per cytoband across samples (rows), ordered by global CNA score (GCS). Sample annotations include age, grade, molecular subtype and local recurrence status at 10 years. **c** Heatmap of twenty genomic regions significantly associated with 10-year risk of local recurrence (Firth-corrected cox model p-value < 0.05 adjusted for grade), showing copy number status. CNAs present in at least 5 patients are shown, with copy number gains in red and losses in blue.

Global CNA burden varied considerably across samples, with approximately half showing minimal alterations (global CNA score < −0.58; **Fig. 6b**). Basal-like tumors exhibited greater CNA burden, while normal-like tumors showed fewer alterations (**Fig. 6b**, **Supplementary Fig. 2a**). Similarly, high-grade lesions showed a greater CNA burden compared to low-grade lesions (**Supplementary Fig. 3b**).

We identified distinct CNA patterns across molecular subtypes: basal-like tumors showed enrichment for gains on 8q, 13q, and 19q; LumA tumors frequently exhibited 16q loss; and Her2-enriched tumors showed characteristic 17q12 gains corresponding to the ERBB2 locus (Fisher’s exact test, p < 0.005; **Supplementary Fig. 3c**). Samples and regions were clustered using Ward’s hierarchical clustering with Minkowski distance metric.

Fourteen cytoband regions were significantly associated with increased 10-year local recurrence risk after grade adjustment (Firth’s penalized likelihood Cox regression p-value & permuted p-values < 0.05; **Fig. 6c**). These included copy number losses in five non-adjacent cytobands and gains in three non-adjacent cytobands. When analyzing invasive local recurrence specifically, four regions were shared (losses in 5q32-33, 9p21, 18p11), with two additional losses identified in 9p13 and 8q11 and one gain in 5p15 (Firth’s penalized likelihood Cox regression p-value < 0.01 & permuted p-value < 0.05).

Copy number losses could alter critical tumor suppressor functions, as evidenced by the presence of several well-known tumor suppressor genes within these regions. Notable examples include *CDKN2A* at 9p21 (a key regulator of cell cycle progression), *PRDM4* at 12q23-24 (involved in cell differentiation and tumorigenesis) [29], *SPARC* at 5q33 (important for extracellular matrix synthesis and modulation of cell shape) [30] and MITOSTATIN at 12q24 (a mitochondrial protein with tumor suppressor activity) [31]. Additional loss highlights disruptions in cell adhesion and intercellular contacts as loss of 18p11 may affect DAL-1, a known regulator of cell adhesion and link between cell membrane and cytoskeleton [32,33].

Gains were identified in regions harboring oncogenes or potential cancer-related genes. For example, amplification in the 11q region—including 11q13, which contains *CCND1* and *CTTN*— previously associated with breast cancer aggressiveness [34,35]. Additionally, a gain in 5p14 includes four cadherin genes (*CDH10*, *CDH9*, *CDH12*, *CDH18*) and *PRDM9*, whose aberrant expression has been associated with genomic instability [36].

Collectively, these findings demonstrate that the CNAs associated with 10 year local recurrence not only affect regions harboring established tumor suppressors and oncogenes but also target regions involved in cell structural integrity and cell-cell interactions, potentially contributing to an aggressive and treatment-resistant phenotype.

Contextualizing our genomic findings with prior profiling studies: consistent early mutational drivers, novel subtype-specific CNA patterns, and predictors of RT response

Most prior DCIS genomic studies analyzed small cohorts (<100 cases) and primarily focused on DCIS cases with concurrent invasive disease (synchronous DCIS) rather than pure DCIS (**Supplementary Table 2**). While recent larger studies, such as those by Strand et al. (2022) [20] and Kader et al. (2024) [37], examined hundreds of pure DCIS cases, their reliance on low-pass sequencing or lack of matched normal tissues limits the sensitivity for detecting genomic changes, particularly when working with FFPE samples, which are often the only available option for DCIS studies.

Across studies, *PIK3CA* and *TP53* consistently emerged as the most frequently mutated genes in DCIS (and invasive disease), with mutation rates ranging from 21–55% and 17–52%, respectively (**Supplementary Tables 2-3**). In our cohort, *PIK3CA* and *TP53* were also among the most frequently mutated genes and identified as tumorigenesis drivers in pure DCIS. Mutations in *GATA3* and *ERBB2,* which were frequently reported in several studies, were only detected in a small number of lesions in our cohort (n = 3 and 7, respectively, **Supplementary Table 3**). Overall, we observe very little overlap between genes reported as frequently mutated across studies (**Supplementary Table 3**). Differences across studies likely reflect variations in methodology, including variant-calling pipelines, sample types (e.g., synchronous vs. pure DCIS), cohort composition (e.g., histological grade and ER/HER2 status), and our study’s specific focus on LR and RT response, complicating direct comparisons. Nevertheless, the consistent identification of *PIK3CA* and *TP53* as the most frequently mutated genes across studies and in our cohort reinforces their pivotal roles in the early stages of breast tumorigenesis.

Recurrent CNAs in regions such as 1q, 8q, and 17q gains, as well as 8p, 11q, and 16q losses, were observed in our study, consistent with prior DCIS findings, including those from Strand et al. (2022) [20] and Abba et al. (2015) [15]. For example, Strand et al. (2022) identified 29 recurrent CNAs in DCIS but found no single CNA predictive of recurrence [20]. In contrast, we identified twenty genomic regions significantly associated with 10-year local recurrence risk, including regions containing known tumor suppressors and oncogenes. These differences may reflect the higher resolution of our sequencing approach (100x whole-exome sequencing) compared to the low-pass sequencing used in prior studies. Our study also provides valuable new insights into molecular subtype-specific CNA patterns in DCIS. In our cohort, basal-like tumors exhibited gains on 8q, 13q, and 19q; Luminal A tumors displayed 16q loss; and Her2-enriched tumors showed 17q12 gains encompassing *ERBB2*. Aside from amplifications in HER2-positive subtypes, subtype-specific CNA analyses remain poorly represented in the DCIS literature but are well established in invasive breast cancer (**see Supplementary Text**). These findings suggest that genetic and molecular aberrations defining subtypes likely arise early and are at least partially established at the DCIS stage.

Finally, while several studies have explored prognostic markers of recurrence, few account for treatment variation, and none specifically examined markers of RT response (**Supplementary Table 2**). Our study uniquely identified genetic alterations within a gene network that integrates cytoskeletal integrity, cell-cell interactions, cell adhesion, and metabolism which are associated with an increased risk of local recurrence within five years following RT. These findings suggest that these alterations may play a critical role in resistance mechanisms activated by RT-induced stress.

## Discussion

Our study provides a comprehensive analysis of pure DCIS, focusing on DNA profiles associated with disease prognosis and response to RT. While previous research has primarily examined genomic changes in synchronous DCIS compared to IDC or pure DCIS, our work offers a detailed view of the mutational landscape specific to pure DCIS and its association with LR. Importantly, we identified genomic alterations associated with molecular subtypes and, most critically, LR—both independent of treatment and specifically in cases following RT— highlighting potential molecular drivers of treatment outcomes. We also observed distinct mutational processes in early-onset DCIS with high tumor mutational burden potentially driven by impaired mismatch repair, but these were not associated with prognosis.

While driver genes like *TP53* and *PIK3CA* are critical in the early stages of tumorigenesis, our study did not find any significant associations between their mutations and disease progression or response to treatment. These findings suggest that TP53 and PIK3CA mutations are essential for overcoming initial biological constraints during early tumor development but may become less relevant to progression once these barriers are surpassed. This aligns with prior research showing that these mutations are frequently retained in invasive carcinoma but occur at higher prevalence in hyperplastic and DCIS lesions [38].

Furthermore, their prognostic relevance appears to depend on tumor clinicopathological characteristics. For example, Lin et al. [11] observed an inverse association between PIK3CA kinase domain mutations in high-grade DCIS tumors and progression, while Silwal-Pandit et al. [39] reported that the prognostic impact of *TP53* mutations varied across molecular subtypes of invasive breast cancer. These findings underscore the complexity of breast cancer progression, where early driver mutations interact with additional genetic and microenvironmental factors to shape disease outcomes.

We identified recurrent CNAs frequently reported in the literature, including gains on 8q [10–12,14,15,37,40], 17q [10,12,15,18–20,37,40], 20q [37,40], and losses on 11q [10–12,18,19,37] and 16q [10,13,14,20,40]. These CNAs mirror those observed in invasive ductal carcinoma, suggesting that pure DCIS already harbors genomic features characteristic of invasive cancer—including alterations that define molecular subtypes (e.g., basal, luminal-A, or Her2-enriched tumors; **see Supplementary Text**). Moreover, this observation is consistent with earlier transcriptional and epigenetic studies that show subtype-specific alterations in gene expression and DNA methylation are already evident in DCIS [41,42]. These findings suggest that genetic and molecular aberrations defining subtypes likely emerge early - being at least partially established at the DCIS stage - and support a model of subtype-specific progression from DCIS to invasive breast cancer [41].

Importantly, our study also revealed novel associations between mutations in genes regulating cytoskeletal integrity, cell membrane organization and function, cell-cell interactions, and cell adhesion, and the risk of LR in pure DCIS. Across the cohort, five genes were significantly associated with LR, underscoring their potential roles in tumor progression. These mutations were not frequently co-occurring in the same patient, and involved genes maintaining cell adhesion (*MYO7A*, *SH2B2*), organizing and supporting cellular organization, cytoskeleton and membrane function (*PDZD8*, *MUCL3*, *MYO7A*). When the cohort was stratified by RT administration, 27 unique genes - excluding *SH2B2* and *MUCL3* - were significantly associated with increased LR risk in RT-treated patients. Many of these mutations were identified in genes critical for maintaining cellular architecture, polarity, and adhesion, such as *KRT1*, *MUCL3*, *TMEM132C*, *TTLL10*, *EML5*, *CEP128*, *CFAP47*, and *KIF24*, highlighting how disruptions in structural integrity and epithelial cell organization may undermine the effectiveness of RT and contribute to LR. In addition to structural changes, metabolic regulators like *MGAM2* and *AADACL3* may influence the energetic and biosynthetic demands required for cell survival and adaptation under genotoxic conditions. Alterations in *REV1*, a key player in DNA damage tolerance, underscore the importance of effective DNA repair mechanisms following RT. These findings expand upon prior studies linking disruptions in tissue structure and cell adhesion to DCIS progression [43–46], while providing novel insights into their specific roles in recurrence following RT in pure DCIS.

Notably, our analysis also revealed distinct biological programs underlying in-situ versus invasive recurrence following RT. Specifically, while some mutations in genes involved in structural maintenance and mechanosensory pathways were uniquely associated with in-situ relapse, invasive progression was characterized by alterations in cell-cycle regulation, genome stability, and inflammatory signaling. Because stratifying by both treatment and LR type reduced our statistical power, these molecular distinctions will require confirmation in larger cohorts.

Finally, we uniquely identified losses in cytobands harboring established tumor suppressor genes—such as *CDKN2A* in 9p21 and *PRDM4* in 12q23–24—as well as regions containing genes crucial for cell adhesion and migration, including a cluster of keratin genes in 12q13–14 and *DAL-1* in 18p11. In parallel, gains in regions like 11q, which includes oncogenic drivers such as *CCND1* and *CTTN*, suggest that oncogene amplification may further contribute to tumor aggressiveness and ultimately patient prognosis. Gains at 5p14, containing cadherin genes (*CDH10*, *CDH9*, *CDH12*, *CDH18*), again emphasize the importance of maintaining epithelial integrity to prevent LR in pure DCIS patients.

Future studies are warranted to investigate how these mutations interact with other molecular pathways and microenvironmental factors to elucidate their contribution to DCIS prognosis and the adverse effects of RT. Such research could help identify strategies to mitigate recurrence risk following RT and improve treatment outcomes [47–49].

## Conclusions

Our findings uncover the genomic landscape of pure DCIS and highlight key factors that contribute to LR and mediate the adverse effects of RT. While *TP53* and *PIK3CA* mutations play important roles in early tumorigenesis, they do not predict recurrence, emphasizing the need for alternative biomarkers. We identify distinct mutational processes and genetic alterations that disrupt cytoskeletal integrity, cell-cell interactions and cell adhesion, potentially destabilizing the epithelial tissue environment and contributing to recurrence, particularly following RT-induced stress. Importantly, our data also suggest that in-situ and invasive recurrences after RT may follow divergent molecular trajectories—mutations affecting structural maintenance and mechanosensory pathways were specifically linked to in-situ relapse, whereas alterations in cell-cycle regulation, genome stability, and inflammatory signaling characterized invasive progression. These insights provide a foundation for understanding the genetic basis of DCIS progression and identifying potential molecular drivers of treatment resistance. Future research will be essential to translate these insights into clinical practice, guiding the development of more targeted therapeutic approaches to improve outcomes for patients with DCIS.

## Methods

### The Ontario DCIS cohort

The Ontario DCIS Cohort was established at the Sunnybrook Health Sciences Center (Toronto, Canada) as a population-based sample of women diagnosed with pure DCIS defined as in situ cancer without any invasive component between 1994 and 2003 [50–52]. All patients underwent BCS, with a subset receiving subsequent RT as part of their standard-of-care.

Adjuvant endocrine therapy was administered to less than 15% of individuals, while none received systemic chemotherapy or neoadjuvant endocrine therapy. The cohort features comprehensive annotation of clinical annotation and expert pathology review. Previous studies of this cohort have characterized outcomes based on clinical factors including age at diagnosis, pathological features (tumor size and nuclear grade), and treatment modalities [53–56].

Sample selection was prioritized to achieve balanced representation between RT-treated and untreated patients, with equivalent distributions of individuals who did or did not develop an invasive or in-situ ipsilateral recurrence within ten years. Tissue cores were obtained from FFPE blocks, sampling DCIS tumors without microinvasion alongside adjacent normal and stromal tissues. DNA and RNA were extracted using the Qiagen AllPrep FFPE DNA/RNA kit (Qiagen). Samples yielding sufficient DNA quantities underwent library construction using the Agilent SureSelect Human Exome library preparation kit and were sequenced on the NovaSeq6000 platform (100bp paired-end, 100M reads/sample) at the Genome Quebec Innovation Centre (Montreal, Canada). While high-quality sequencing data was obtained for 300 tumor tissues, downstream analyses focused on 147 samples with matched normal profiles (144 normal tissue and 3 stroma non-epithelial samples).

Molecular subtypes were determined using RNA profiles available for a subset of patients (n = 122). Sequencing libraries were prepared using RNA Flex kit (Illumina). Raw reads were processed using `TrimGalore` to remove adapters and low-quality reads [57], followed by alignment to the human genome (Ensembl release v104) using `STAR` [58]. Reads counts were mapped to genomic features using `Rsubread` R package [59] and gene counts were normalized using the variance stabilizing transformation implemented in the `DESeq2` R package [60]. PAM50 subtype classification was performed using the `genefu` R package [61] with established centroids [62]. Specifically, normalized expression data of the 50 PAM50 genes were obtained using variance stabilizing transformation implemented in the `DESeq2` R package [60] and compared to subtype-specific centroids using Pearson correlations. Each sample was assigned to the molecular subtype with which it showed the highest correlation coefficient.

### Whole-exome sequencing data preprocessing

Raw reads were processed using Trimmomatic [63] (version 0.39) to remove adaptor sequences and low-quality bases. Reads were trimmed to retain high-quality sequences, applying quality thresholds of 10 at read ends and 20 within a 4-base sliding window. The remaining paired reads were processed following the GATK4 best practices. Briefly, reads were aligned to the human reference genome (GRCh38, GATK resource bundle) using the

Burrows-Wheeler Aligner (BWA) [64]. Post-alignment procedures included sorting, annotating reads with read groups, and marking duplicate reads with Picard. Base quality score recalibration was conducted using GATK4 tools. A recalibration table was generated with the `BaseRecalibrator` function using known variant sites (dbSNP138 for SNPs and Mills and 1000 Genomes for indels), and recalibration was applied with `ApplyBQSR` to adjust base quality scores and correct for systematic technical errors. The process focused on SureSelect

Human Exons v7 regions with a 100 bp padding. Finally, properly paired reads were extracted, excluding secondary alignments and low-quality reads, with the resulting files indexed using `Samtools` [65]. After filtration, samples had a mean coverage depth of 152X (range: 52-308X, standard deviation: 42X).

### Single nucleotide variant & indel calling

For variant calling, we used NeuSomatic [66], a deep learning approach that leverages both tumor and matched normal sequence alignment information, alongside somatic mutation calls from six different approaches: MuTect2, MuSE, VarDict, VarScan2, Strelka2, and SomaticSniper [67–72]. This method was selected because of the low level of agreement between callers in our data - the majority of mutations (88.7%) were identified by only one caller (**Supplementary Fig. 4a**) - an observation consistent with previous studies [73–75]. The number of mutations detected varied significantly across samples, with the minimum identified in any single sample being 1,563 mutations, and the maximum reaching 283,247 mutations (**Supplementary Fig. 4a**).

Small-variant calling has become increasingly amenable to deep learning approaches, thanks to the availability of extensive sequencing data and robust benchmarking datasets that cover millions of variants across diverse genomic contexts [76]. A convolutional neural network (CNN) architecture leverages information from sequencing reads and the reference genome in the vicinity of each candidate variant to approximate complex, nonlinear functions, accurately classifying loci as homozygous variant, heterozygous variant or homozygous reference (non-variant) [66].

Best practices established using well-characterized somatic reference datasets from the SEQC-II consortium demonstrated that NeuSomatic models achieve robust performance across various sequencing technologies - including both fresh and FFPE DNA inputs - across a range of tumor/normal purities and sequencing coverages, significantly outperforming conventional approaches [73]. Accordingly, we used the ensemble extension of NeuSomatic. This extension combines outputs from six individual variant callers by integrating features from 93 channels and incorporates an additional 26 channels to capture alignment information in a window of seven bases around the candidate mutation, resulting in a total of 119 input channels per candidate matrix. We used the recommended pre-trained model SEQC- II (SEQC-WGS-Spike model), which was trained on 20 whole-genome sequencing replicate pairs containing *in silico* somatic mutations with allele frequencies ranging from 1% to 100% (with matched normals at both 95% and 100% purity, and sequencing coverage ranging from ∼40x to 220x) using five variant callers used: MuTect2, Strelka2, MuSE, SomaticSniper, VarDict. After mutation calling, the recommended post-processing was applied to resolve long

INDEL sequences and the final NeuSomatic predictions were used for all downstream analyses.

We obtained comprehensive genomic information for each variant using the Ensembl Variant Effect Predictor (VEP) [77]. This included the effects on gene and protein function, such as consequence types and amino acid changes, variant frequencies in different populations, impact on regulatory regions, and potential associations with diseases and phenotypes.

Following annotation with the Ensembl VEP, we used the `vcf2maf` tool to transform VEP-annotated VCF files into the Mutation Annotation Format (MAF). This conversion ensures each variant is uniquely associated with a single gene transcript or isoform, despite the potential for a variant to impact multiple isoforms. Particularly in cases where variants could be classified under different effects — such as a `Missense_Mutation` near a `Splice_Site` — the MAF format forces a singular designation for each variant by leveraging VEP’s determinations for canonical isoforms.

We excluded 100 genes commonly mutated in public exome datasets (FLAGS) due to their lower likelihood of disease association [78]. This decision stems from their longer coding regions, which inherently increase mutation probability, and the presence of paralogs that might offset functional loss these mutations could cause [78,79].

High-confidence variants (identified with a probability score of 0.7 or higher) consistently showed higher allele frequencies compared to those categorized as low-quality (with scores between 0.4 and 0.7) and rejected variants (with scores below 0.4) (**Supplementary Fig. 4B**). To reduce potential false positives, we selected high-confidence variants with allele frequency above 0.1. To further confirm that variants detected were not technical artifacts, we assessed the relationship between tumor mutational burden and sequencing depth (**Supplementary Fig. 5**). The lack of correlation supports robust variant detection independent of coverage.

Mutation patterns and frequencies were visualized using the oncoplot function from the maftools R package [80], which displays mutation types and frequencies across samples.

### Mutational signatures

We performed mutational signature analysis using the COSMIC database of single-base substitution (SBS) signatures [24]. First, the trinucleotide context of each single nucleotide variant was characterized. We then used the `fit_to_signature` function in the `MutationalPatterns` R package (version 3.19) [81] to find the linear combination of COSMIC mutation signatures that most closely reconstructs the mutation spectra for each sample by solving the nonnegative least-squares constraints problem. We used strict refit where the signature with the lowest contribution is removed; refitting is repeated until the cosine similarity between the original and reconstructed profile becomes more than max_delta= 0.004). We selected signatures that contributed to the mutation spectrum of at least 10 samples and plotted their relative contributions using the `pheatmap` R package [82].

### Driver genes

To identify driver genes (i.e., genes under positive selection in cancer), we used the `dNdScv` analysis method [26]. This approach is based on the evaluation of the ratio between synonymous (silent) mutations and non-synonymous (missense) mutations in genes. A higher ratio of non-synonymous to synonymous mutations in a gene indicates positive selection for mutations that may confer a growth advantage to cancer cells, suggesting the potential role of the gene as a driver in tumorigenesis.

The `dNdScv` method estimates the background mutation rate of each gene by combining local information (synonymous mutations within the gene) with global information (variation of mutation rates across genes). This approach controls for the sequence composition of genes and accounts for mutational signatures, providing a more accurate estimation of the expected neutral mutation rate. In particular, the `dNdScv` R package [83] implementation uses trinucleotide context-dependent substitution matrices to mitigate common mutation biases that can affect dN/dS calculations [26].

To visualize and analyze the distribution and nature of mutations in driver genes, we used lollipop plots generated from the maftools R package [80] and annotation tracks from cBioPortal [84]. These plots provide a representation of mutation types and their locations along the protein sequence as well as annotations including likely mutation hotspots as identified by Memorial Sloan Kettering Cancer Hotspots and 3D Hotspots databases [85], and annotation records of therapeutic indication from OncoKB [86].

### Copy Number Alterations (CNAs)

To investigate copy number alterations, we applied the Allele-Specific Copy Number Analysis of Tumours (ASCAT v3) on our tumor normal pairs estimating tumor purity, ploidy, and allele-specific copy number [87]. The `runAscat` function was executed with default settings optimized for high-throughput exome sequencing data, with the gamma parameter set to 1.

After examination of ASCAT sunrise plots, we identified a subset of samples (n=35 samples), for which the initial estimates of tumor purity and ploidy did not align with the regions of highest confidence on the sunrise plots. The `runAscat` was re-run for these samples by manually assigning the aberrant cell fraction (tumor purity) and tumor ploidy parameters corresponding to the regions of highest probability as depicted on the sunrise plots. Overall, eight samples were excluded from further analysis due to poor goodness of fit leaving 139 samples with CNA profiles for downstream analyses. Absolute number gains and losses shared across samples were visualized across whole chromosomal regions using `aCNViewer` [88] (window size of 1 Mbp). The estimated ploidy for each sample is subtracted from the copy number values of each segment which means that a copy number of 0 is copy number change. These adjusted windows at base resolution are then plotted into a stacked histogram representing genome-wide absolute copy number and copy neutral variations over all samples in a group.

We applied a re-segmentation approach to adjust for amplitude divergence due to technical variability implemented in `CNApp` [89] using the default settings (minimum segment length = 100 Kbp, minimum amplitude deviation from segment to zero = 0.16, maximum distance between segments=1 Mb, maximum amplitude deviation between segments = 0.16, and maximum BAF deviation between segments = 0.1). Re-segmented data were then used to calculate the broad, focal and global CNA scores. We then transformed re-segmented data into genomic regions profiles (chromosome arms, cytobands and sub-cytobands) using both focal and broad segments. Length-relative means are computed for each window by considering amplitude values from those segments included in each specific window. Default cutoffs for low-level copy number gains and losses (i.e., |0.2|) were used to infer CNA frequencies.

### Survival analyses

We evaluated the association between gene mutations or copy number aberrations in cytobands and 10-year local recurrence-free survival using Firth’s penalized likelihood Cox regression which accounts for small sample sizes and rare events. This analysis was conducted using the `coxphf` R package [90]. Aberrations were included in the analysis only if detected in at least five lesions (2,030 genes; 303 cytobands). This analysis was performed across the entire patient cohort. To further investigate mutated genes associated with response to RT, a stratified analysis was conducted based on treatment groups.

To control for multiple testing in the identification of mutated genes associated with LR, we employed permutation-based testing (1,000 permutations) to establish empirical significance thresholds. We required both a traditional p-value threshold (p < 0.01 for full-cohort analyses; p < 0.05 for RT-stratified analyses) and a permuted p-value < 0.05.

Kaplan-Meier survival curves were used to visualize the results, illustrating event-free survival probabilities over time for patients stratified by mutational status in the specified genes or gene sets.

GO enrichment analyses were performed using the clusterProfiler R package [91]. Entrez gene identifiers were mapped to GO terms using the `org.Hs.eg.db` annotation database [92]. All GO terms with FDR < 0.2 were considered. Semantic similarity between GO terms was calculated using the Wang method implemented in the `pairwise_termsim` function [93]. The enrichment map was visualized using the `emapplot` function [94] which displays the significantly enriched terms with at least 2 genes.

### Third-party studies

A systematic literature search was performed using PubMed to identify previous studies that conducted DNA profiling on pure DCIS or DCIS mixed with invasive lesions. The search strategy included terms related to “ductal carcinoma in situ”, “genetic markers”, “DCIS prognosis”, “DCIS progression”, “DCIS to IDC”, “dcis dna”, “dcis prognosis dna markers”, “copy number alterations”, and “somatic mutations”. Studies were included if they reported genomic analyses of DCIS samples using sequencing or copy number profiling techniques and were published within the last 10 years. Twelve studies met the inclusion criteria, and their key findings were summarized in **Supplementary Table 1**. The review emphasized genetic alterations and pathway dysregulation that may drive DCIS initiation and progression to invasive disease.

## Declarations

### Ethics approval and consent to participate

Research ethics board approval was obtained from Sunnybrook Health Sciences Centre, Toronto, Ontario (#2738). This study was facilitated through ICES which is named as a prescribed entity in section 45 of PHIPA (Regulation 329/04, section 18) which allows access and utilization of administrative data for research purposes with a waived requirement for consent.

### Consent for publication

Not Applicable

### Availability of data and materials

The dataset analyzed in this study is not publicly available due the personal, sensitive, and inherently identifying nature of raw genomic data. Access to raw data and patient metadata may be provided but is controlled and requires institutional material data transfer agreements (contact person).

The scripts used to reproduce the analyses performed in this study are available in the Dumeaux Lab GitHub Repository https://github.com/dumeaux-lab/dcis-dna_paper.

## Funding

This work was supported by Canadian Institute for Health Research (CIHR) project grant #391682. We thank the Canadian Foundation for Innovation J. Evans Leaders Fund grant #43481 for the support of the computing infrastructure used throughout our analyses.

ER holds LC Campbell Chair for breast cancer research. NR received Breast Cancer Canada and Ontario Graduate Scholarships.

We thank Genome Quebec Innovation Center for RNA processing and sequencing.

## Data Availability

Owing to the personal, sensitive and inherently identifying nature of raw genomic data, access to raw data and patient metadata is controlled and requires institutional material data transfer agreements (contact person).

https://github.com/dumeaux-lab/dcis-dna_paper

## Data Acknowledgement

This study was supported by ICES, which is funded by an annual grant from the Ontario Ministry of Health (MOH) and the Ministry of Long-Term Care (MLTC). The opinions, results and conclusions reported in this paper are those of the authors and are independent from the funding sources. No endorsement by ICES, the MOH or MLTC is intended or should be inferred. As a prescribed entity under Ontario’s privacy legislation, ICES is authorized to collect and use health care data for the purposes of health system analysis, evaluation and decision support. Secure access to these data is governed by policies and procedures that are approved by the Information and Privacy Commissioner of Ontario.

## Authors’ Contributions

Conceptualization, S.N.M, L.P., E.R., M.T.H and V.D.; Methodology, N.R, E.S, V.D.; Investigation, N.R., E.S., M.Z., V.D.; Software N.R., E.J.M., V.D.; Writing – Original Draft, N.R. and V.D.; Writing – Review & Editing, E.S., M.Z., E.J.M., E.H., S.N.M., S.T., L.P., M.T.H. and E.R. ; Funding Acquisition, E.R., M.T.H, E.H., S.N.M, S.T., L.P. and V.D.; Resources, E.R., S.N.M, S.T., L.P. ; Supervision, V.D.

