## Supplementary Information for "Mutational landscape of pure ductal carcinoma in situ and associations with disease prognosis and response to radiotherapy"

### Supplementary Text: Comparison to invasive breast cancer subtypes

When compared to invasive breast cancer subtypes [1], our findings reveal both similarities and differences in CNA patterns. Invasive basal-like tumors frequently exhibit gains on 8q, 10p, and 12p, as well as losses on 5q and 9p. Although these alterations were observed in our cohort, only 8q gain was significantly enriched in basal-like DCIS. Interestingly, 16q loss, a hallmark of certain genomic-driven IDC subtypes (e.g., IntClust 7 and 8), was also enriched in Luminal A DCIS in our study. IntClust 8, predominantly associated with Luminal A invasive cancers, is characterized by the classical 1q gain/16q loss event, resulting from an unbalanced translocation [1–3] . In contrast, IntClust 7 retains 16q loss but lacks the characteristic 1q gain.

The IntClust 8 subtype also demonstrates high levels of *PIK3CA, GATA3,* and *MAP2K4* mutations, while IntClust 7 is characterized by the highest frequency of *MAP3K1* and *CTCF* mutations across all clusters. These findings suggest that certain molecular features of invasive breast cancer are already present at the DCIS stage, supporting the hypothesis of early divergence in the evolutionary trajectories of different breast cancer subtypes.

**Supplementary Table 1:** Patient and tumor clinical attributes according to radiotherapy (RT) administration

| | | Without RT (n = 74) | With RT (n = 73) | p-value |
|---|---|---|---|---|
| Local Recurrence N (%) | DCIS | 15 (10.2%) | 13 (8.8%) | 0.48 |
| | Invasive | 25 (17.0%) | 19 (12.9%) | |
| | None | 34 (23.1%) | 41 (27.9%) | |
| Time to LR (years) Mean (Range) | DCIS | 3.4 (0.5 - 9.7) | 2.7 (0.9 - 8.1) | 0.54 |
| | Invasive | 4.0 (0.6 - 9.2) | 5.0 (1.7 - 9.7) | 0.34 |
| Clear Margins N (%) | Positive | 7 (4.8%) | 3 (2.0%) | 0.29 |
| | Negative | 60 (40.8%) | 59 (40.1%) | |
| | Undetermined | 7 (4.8%) | 11 (7.5%) | |
| Tumor size (mm), Median (Range) | | 18.5 (2.0 - 90.0) | 17.5 (4.0 - 55.0) | 0.63 |
| Nuclear Grade N (%) | Low | 3 (2.0%) | 1 (0.7%) | 0.58 |
| | Moderate | 44 (29.9%) | 43 (29.2%) | |
| | High | 27 (18.4%) | 29 (19.7%) | |
| Multifocality N (%) | Present | 18 (12.2%) | 18 (12.2 %) | 0.57 |
| | Absent | 39 (26.5%) | 33 (22.4%) | |
| | Undetermined | 17 (11.6%) | 22 (15.0%) | |

| | | | | |
|---|---|---|---|---|
| Age<br>N (%) | <50 Years Old | 23 (15.6%) | 29 (19.7%) | 0.03 |
| | 50-60 Years Old | 20 (13.6%) | 28 (19.0%) | |
| | >60 Years Old | 31 (21.1%) | 16 (10.9%) | |
| PAM50 Subtype (Pearson Correction)<br>N (%) | Basal-like | 14 (9.5%) | 7 (4.8%) | 0.23 |
| | Her2-like | 6 (4.1%) | 12 (8.2%) | |
| | LumA | 15 (10.2%) | 17 (11.6%) | |
| | LumB | 13 (8.8%) | 8 (5.4%) | |
| | Normal-Like | 12 (8.2%) | 13 (8.8%) | |

**Supplementary Table 2:** Review of DCIS genomic studies published in the past ten years

| **Dataset** | **Groups** | **#** | **Mean follow-up (years)** | **Profiling** | **Variant calling** | **Clinico-pathological characteristics** | **RT treatment** | **Main finding** |
|---|---|---|---|---|---|---|---|---|
| Lin et al. 2019 [4] | Pure DCIS | 65 | 13 | Targeted exon sequencing of most prevalent mutations identified in TCGA IBC (223 genes/regions) | Identified by at least 2: MuTect, VarScan2, VarDict, or Freebayes | Mostly moderate to high grade, Her2-enriched or LumA. half had mastectomy | Unknown | - Most prevalent PIK3CA (34.4%) and TP53 (18.4%)<br>- No significant difference of mutational burdens in selected genes between the two groups of DCIS cases<br>- Inverse association between *PIK3CA* kinase domain mutations and progression<br>- Copy number variations in 1q32 and 8q24 associated with progression |
| | Synchronous DCIS | 60 | | | | | | |
| Hernandez et al. | Synchronous DCIS | 13 | NA | Sequenom MassARRAY, | N/A | Mostly low to moderate grade, ER+, PR+. Fresh | Unknown | - *PIK3CA* mutant allele present |

| | | | | | | | | |
|---|---|---|---|---|---|---|---|---|
| 2012 [5] | | | | using OncoCarta Panel v1.0, (targets 238 mutations across 19 oncogenes), followed by<br>Sanger Sequencing | | frozen samples | | at 49% in DCIS<br>- Most frequently amplified genomic regions (three out of 13 cases) mapped to 17q12, encompassing *HER2*, and 11q3-q14, encompassing *CCND1*<br>- Out of all genes tested, only *PIK3CA* was found to be somatically mutated in the samples investigated<br>- The H1047R *PIK3CA* mutation, detected at low frequency in DCIS and absent in the invasive component in 15% of cases |
| Nagasawa et al. 2021 [6] | Pure DCIS | 21 + 72 | 6.1 | Whole Exome (discovery cohort)<br><br>Targeted Sequencing (custom sequencing panel containing 180 genes)(validation) | Mutect2, Genomon pipeline(D) | Mostly ER+, most received breast conserving surgery | Unknown | - *GATA3* and *PIK3CA* mutations were the most commonly detected mutations present in four (19%) and five patients (24%), respectively<br>- Two of the four patients with *GATA3* mutations harbored the same mutation in their primary and relapsed tumors<br>- No patient with PIK3CA mutation experienced relapse, whereas 9 of 16 patients without PIK3CA mutation (56.2%) experienced relapses |
| | Pure DCIS and invasive recurrence | 4 | | | | | | |
| Kim et al. 2015 [7] | Pure DCIS | 6 | NA | Whole Exome | Mutect, SomaticIndelDetector | Mostly pre menopausal, ER+, PR+ | Unknown | -CNA reveals 18 candidate chromothripsis occuring most frequently on chromosomes 8, 17 and 21<br>-Pure DCIS driver genes include *PIK3CA* and *AKT1*<br>-Genes commonly mutated in both pure DCIS and DCIS-IDC included TP53, PIK3CA, CBFB and MAML2<br>- oncogenes PIK3CA, FGFR2 and GNAS involved both somatic |
| | Synchronous DCIS | 5 | | | | | | |

| | | | | | | | | |
|---|---|---|---|---|---|---|---|---|
| | | | | | | | | mutations and copy number gains<br>- tumor suppressor genes TP53, PTEN, BRCA2 and ATM involved both somatic mutations and copy number losses. |
| Nachmanson et al. 2022 [8] | Pure DCIS | 40 | NA | Whole exome and whole transcriptome | Must be identified by both VarDictJava, Mutect2 | Mostly moderate to high grade, ER+, ERBB2 overexpression | Unknown | -Aging-associated mutational signatures (SBS1 and SBS5) in all samples eligible for analysis (N = 13),<br>-mismatch repair signature (SBS15 or SBS21) in three DCIS of variable grade and architecture<br>-The most recurrently mutated genes were PIK3CA (44%), TP53 (31%), and GATA3 (20%)<br>-loss of 16q (13/30) and 17p (12/30) or gain of 1q (12/30) were among the most frequent chromosomal alterations |
| | Recurrent DCIS | 3 | | | | | | |
| Pareja et al. 2020 [9] | Synchronous DCIS | 25 | NA | Whole exome, targeted MSK-IMPACT (≥410 key cancer-related genes) | Whole exome: SNV by MuTect; indels by Strelka and VarScan2 | Moderate to high grade, mostly ER+, Her2- | Unknown | Frequently mutated cancer genes overlapped significantly between DCIS and IDC-NST, including TP53(52%, 54%), PIK3CA (41%, 42%), and GATA3 (26%, 23%).<br>-The majority of synchronous DCIS (63%, 12/19) and IDC-NSTs (58%, 11/19) displayed a dominant aging signature (signatures 1 or 5)<br>-synchronous DCIS (n=27) and IDC-NSTs (n=26) displayed largely comparable copy number profiles, including recurrent 1q and 16p gains, and losses of 5q, 6q, 8p, and chromosomes 13 and 22 |
| | IDC | 26 | | | | | | |
| | Pure DCIS | 7 | | | | | | |
| Abba et al. 2015 [10] | Pure DCIS | 29 | NA | Exome Capture Sequencing | MuTect | High grade (HG) only | Unknown | HG-DCIS cases (62%) displayed mutations affecting one or combinations of targets (driver |

| | | | | | | | | |
|---|---|---|---|---|---|---|---|---|
| | | | | (23,000 genes), | | | | genes) PIK3CA (21%), TP53 (17%), GATA3 (7%), MLL3 (7%)<br>-83% (24 of 29) of DCIS lesions displayed evidence of large chromosomal alteration<br>-The most common regions of increased DNA copy number include chr1q, chr8q, chr17q, and chr20q and regions of common copy number loss include regions chr8p, chr11q, chr17p, and chr22q |
| Kader et al, 2024 [11] | Non-recurrent DCIS | 32 | 8.5 | Targeted sequencing,<br>Whole exome,<br>Low coverage whole genome<br>No matched normal | GATK Unified Genotyper, Platypus, and Varscan 2 | Mean age of 60, ER and HER2 negative | Mostly no (80%) | -Four chromosomal changes (5q, 11q, 17q and 20q) and TP53 mutation were enriched in clonal primaries compared with non-recurrent DCIS<br>-Thirty-seven clonally related cases shared somatic driver mutations, most commonly in TP53 (n=24) and PIK3CA (n=16) |
| | Pure DCIS and in-situ or invasive recurrence | 54 | | | | | | |
| Strand et al, 2023 [12] | Non-recurrent DCIS | 163 | 7.3 | Light-pass whole genome sequencing | none | Mostly High grade | Yes | - CNA analyses, no SNV analyses<br>- 29 recurrent CNAs, none of which are predictive of recurrence<br>-13 gains (most frequent being 1q, 17q) and 16 losses (most frequently noted in 17p, 16q, 11q) occur in 10.1% - 52.6% of DCIS samples<br>-11 hallmark pathways significantly associated with early recurrence (including MYC, mTOR signaling, and cell cycle pathways) |
| | Pure DCIS and in-situ recurrence | 81 | | | | | | |
| | Pure DCIS and invasive recurrence | 69 | | | | | | |
| Kaplan et al, 2024 [13] | Synchronous DCIS | 50 | Unknown | DNA-seq, whole transcriptome, RNA-seq | Tempus xT informatics pipeline | Mostly ER, PR positive and *ERBB2* negative | Unknown | -most frequent genomic variations in *PIK3CA, TP53, KMT2C, MAP3K1, GATA3, SF3B1*<br>-most frequent copy number |

| | | | | | | | | |
|---|---|---|---|---|---|---|---|---|
| | | | | | | | | variants seen in *MCL1, CKSB1 and ERBB2*<br>-ERBB2 changes were not seen in IBC unless present in the corresponding DCIS<br>-top genes associated with DCIS were related to muscle regulation, olfactory receptors, and immune related functions |
| Trinh et al, 2021 [14] | Pure DCIS with invasive recurrence | 6 | Unknown | Whole exome sequencing, RNA seq, cyclic immunofluorescence + DAPI staining | MuTect1, MuTect2, and Strelka | 3 ER+, 2 HER2+, and 1 TN tumors | Unknown | - Frequently observed changes included 1q, 8q, 16p, 17q amplification and 11q and 16q loss<br>- Increase of TILs in DCIS was shown to be associated with both activated and immunosuppressive gene signatures<br>- ER− tumors are more immune hot compared with ER+ tumors<br>- RNA-expressed neoantigens included CDH1, PIK3CA, which were the same hotspot driver mutations preserved in the DCIS-to-IDC transition<br>- TP53 mutation, genomic instability, and telomere crisis are more common in ER− DCIS and have been associated with higher TILs |
| Pang et al 2017 [15] | Pure DCIS | 40 | Unknown | Whole genome sequencing | Agilent SureCall | 16 cases were estrogen receptor (ER)-positive, 14 progesterone receptor (PR)-positive, and 5 cases were HER2 amplified | Unknown | - Frequently mutated genes: PIK3CA (55%), TP53 (30%), and GATA3 (45%)<br>- TP53 mutations were exclusive to high grade DCIS and more frequent in PR-negative tumors<br>- RUNX1 mutations and MAP2K4 loss were novel findings<br>- Frequent CNAs: gains on 1q, 8q, 17q, and 20q and losses on 8p, 11q, 16q, and 17p |

| | | | | | | | | |
|---|---|---|---|---|---|---|---|---|
| Lips et al [16] | Primary DCIS | 24 (WES) + 71 (copy number) | 9.4 | Whole exome sequencing (24 DCIS-INV recurrence pairs), SNParray or low pass WGS (71 DCIS-INV pairs), single-cell sequencing (4 DCIS-recurrence) | Platypus v.0.5.2, Torrent Variant Caller (TVC) v.5.6 | Larger cohort (129 primary DCIS): 52% high grade, 67% ER+, 29% HER2+ in primary DCIS samples | Larger cohort (129 primary DCIS): 13% (12/95) of primary DCIS that developed into IDC received RT, but 53% (18/34) of primary DCIS that developed into pure DCIS received RT | - Most IBC is clonally related to DCIS, except for 18% which are de novo<br>- Non-clonally related cases could be due to genetic disposition, because the same genes also predispose DCIS<br>- TP53, PIK3CA mutations and HER2 amplifications are present at DCIS stage before becoming IBC<br>- Most common amplicons: 17q12, 17q21, 11q13<br>- 1q and 8p11 gains are common in recurrent invasive disease, while 3p21 loss is more common in primary DCIS |
| | Invasive recurrence | 24 (WES) + 71 (copy number) | | | | | | |
| | In-situ recurrence | 1 (single-cell) | | | | | | |

**Supplementary Table 3:** List of genes identified as mutated in DCIS and/or invasive disease in the studies listed in Supplementary Table 2

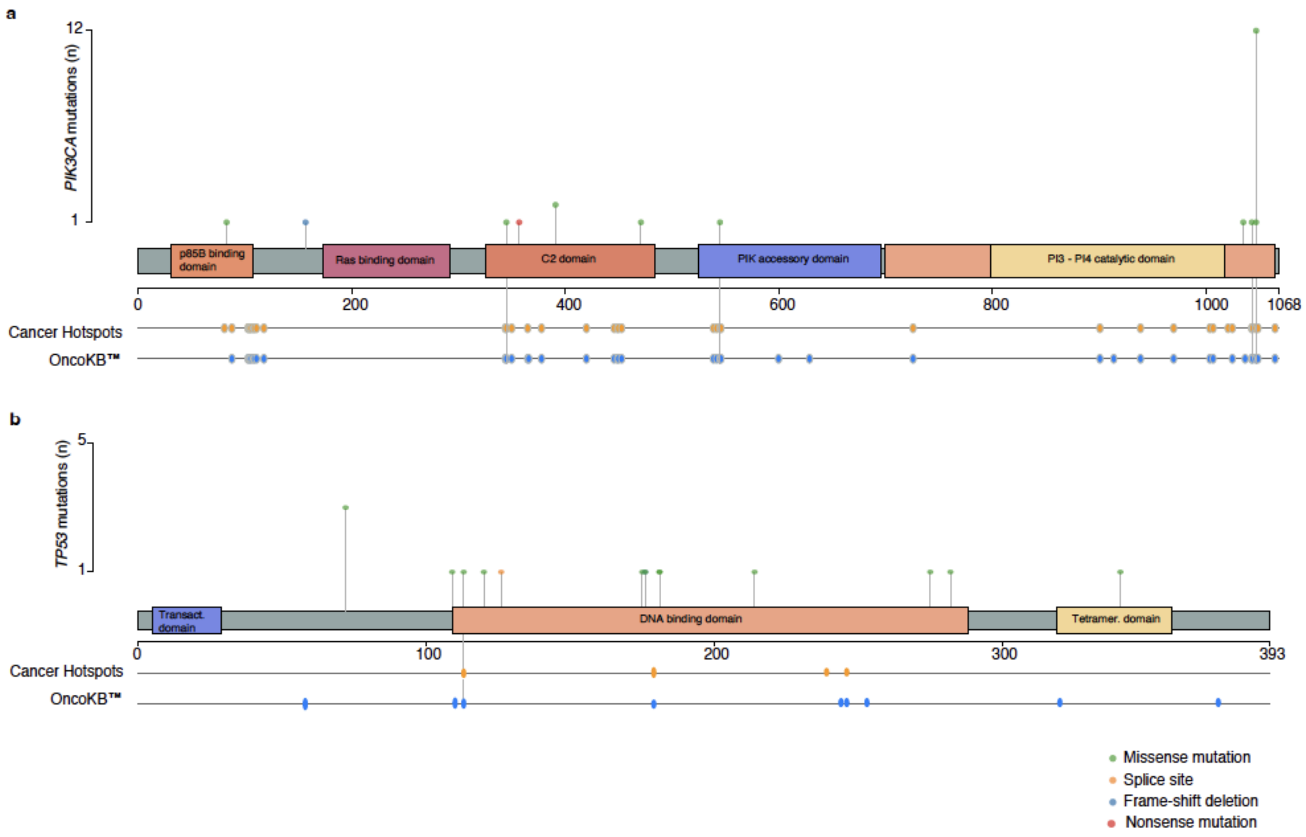

**Supplementary Fig. 1: Distribution of genetic variants identified within canonical transcript of significant driver genes.**

The horizontal axis represents the nucleotide positions along each gene sequence, and the height of each data point corresponds to the count of occurrences of the variant within the cohort. Variants are color-coded based on their mutation type and annotation tracks including cancer hotspot and oncoKB annotations are depicted below each gene sequence for **a** *PIK3CA* and **b** *TP53*.

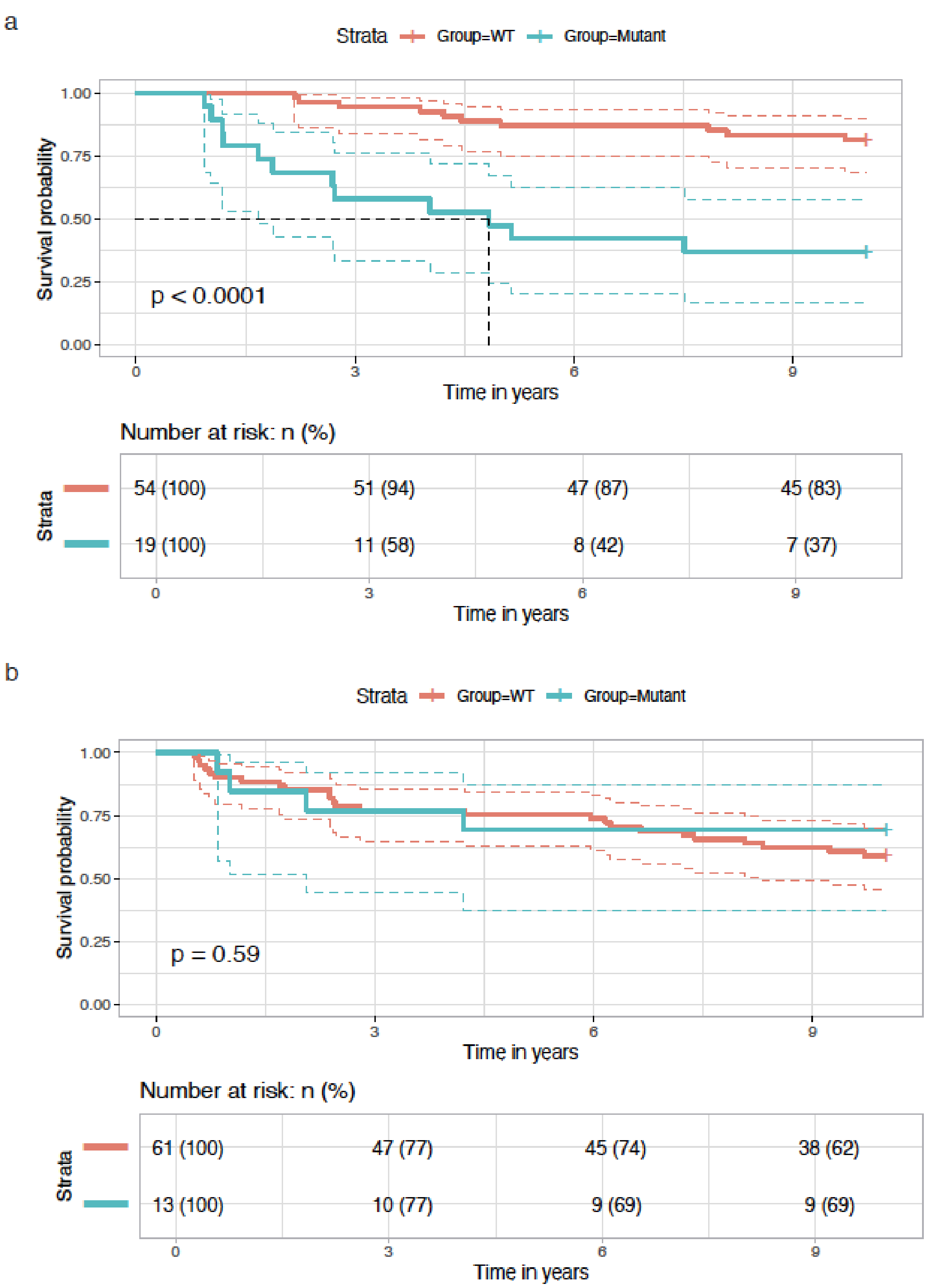

**Supplementary Fig. 2:** KKaplan–Meier analysis of local recurrence–free survival (LRFS) comparing patients with mutations in at least two of a selected panel of 26 genes (red) versus those without such mutations (blue): (a) among RT-treated patients and (b) among non-RT patients. The 26 genes were identified by univariate analysis as being associated with prognosis in RT-treated patients (p-value and permuted p-value < 0.05), after excluding *SH2B2* and *MUCL3* (previously associated with prognosis in the whole cohort) and *ZNF641* (mutated in fewer than 5 lesions in the non-RT group).

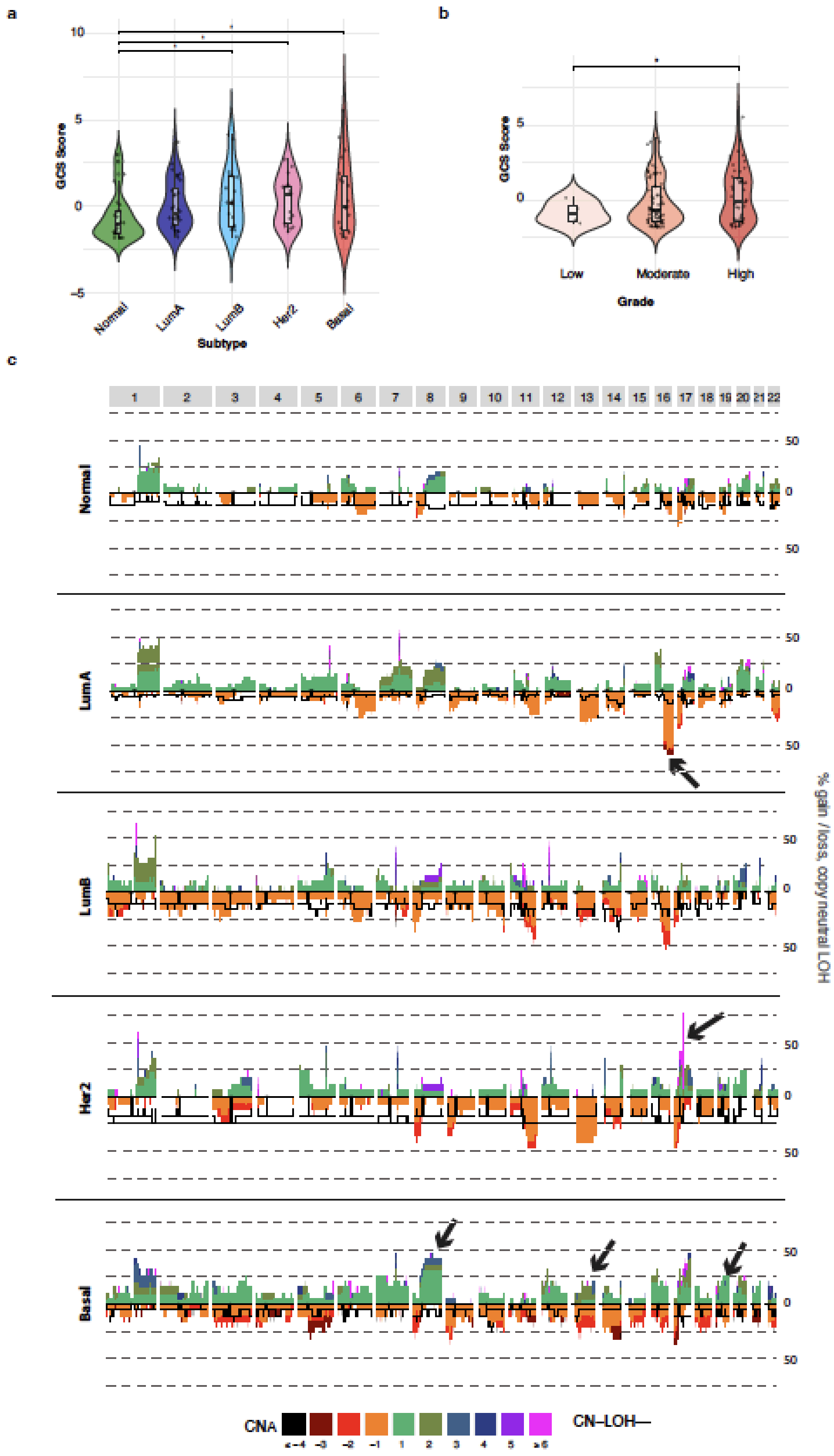

**Supplementary Fig. 3: Global and subtype specific copy number alterations across DCIS lesions.**

**a-b** Distribution of global CNA scores (GCS) across (**a**) molecular subtypes and (**b**) tumor grade. Asterisks indicate significant differences between groups. **c** Frequency plot showing percentage of samples with gains, losses and copy-neutral loss of heterozygosity (LOH) across chromosomes 1-22 for each molecular subtype. Subtype-specific gains and losses are indicated by black arrows.

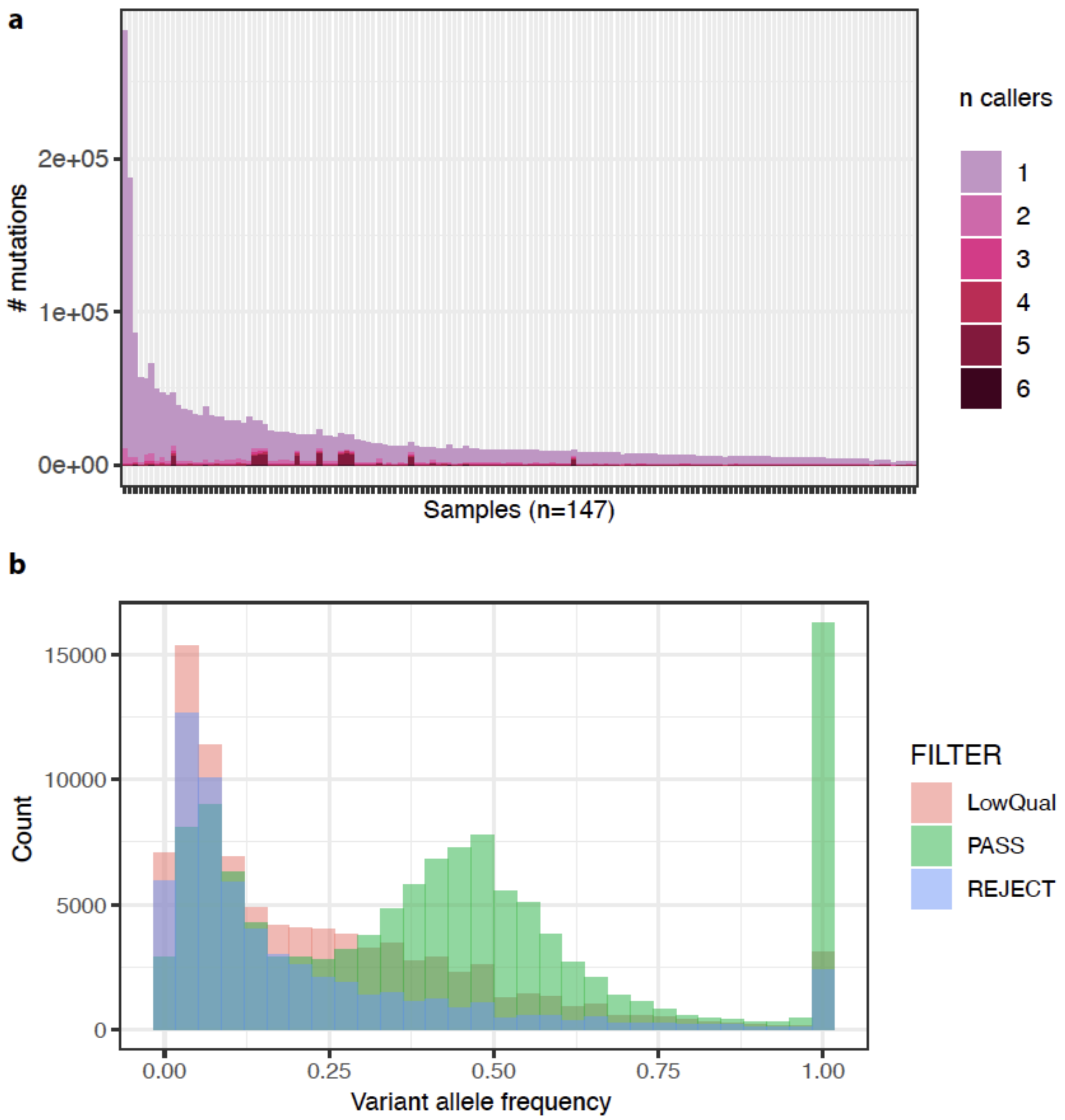

**Supplementary Fig. 4: Single Nucleotide Variant & Indel Calling.**
**a** Caller Concordance in Mutation Detection. Each color indicates the number of mutation callers that have identified the same variant, ranging from unique detections by a single caller to unanimous identifications by all six callers: MuTect2, MuSE, VarDict, VarScan2, Strelka2, and SomaticSniper. **b** Variant allele frequency according to variant confidence score category.

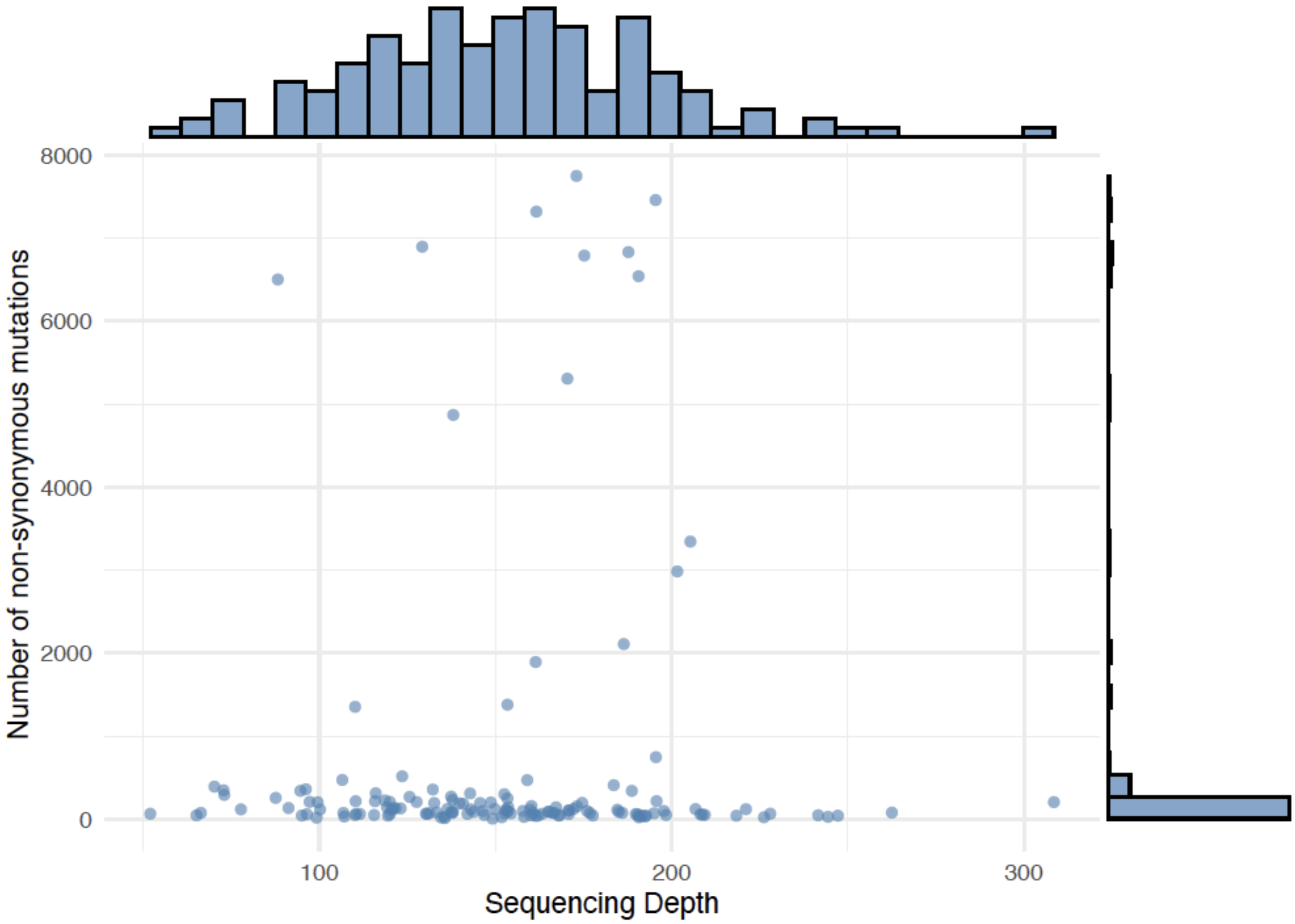

**Supplementary Fig. 5: Number of non-synonymous variants according to sequencing depth**

14. Trinh A, Gil Del Alcazar CR, Shukla SA, Chin K, Chang YH, Thibault G, et al. Genomic Alterations during the In Situ to Invasive Ductal Breast Carcinoma Transition Shaped by the Immune System. Mol

Cancer Res. 2021;19:623–35.

15. Pang J-MB, Savas P, Fellowes AP, Mir Arnau G, Kader T, Vedururu R, et al. Breast ductal carcinoma in situ carry mutational driver events representative of invasive breast cancer. Mod Pathol. 2017;30:952–63.

16. Lips EH, Kumar T, Megalios A, Visser LL, Sheinman M, Fortunato A, et al. Genomic analysis defines clonal relationships of ductal carcinoma in situ and recurrent invasive breast cancer. Nat Genet. 2022;54:850–60.
