## Supplementary material for "Mutational landscape of pure ductal carcinoma in situ and associations with disease prognosis and response to radiotherapy": Table1

|  |  | **Without Local Recurrence (n = 96)** | **With Local Recurrence (n = 51)** | **p-value^a^** |
| --- | --- | --- | --- | --- |
| Type of 10-year Local Recurrence | DCIS | 0 | 24 (47.0%) |  |
|  | Invasive | 0 | 27 (53.0%) |  |
| Time to Recurrence in years  Median (Range) | DCIS | NA | 2.1 (0.5 – 9.7) |  |
|  | Invasive | NA | 4.2 (0.6 – 9.7) |  |
| Radiotherapy (RT)  N (%) | Yes | 51 (53.1%) | 22 (43.1%) | 0.32 |
|  | No | 45 (46.9%) | 29 (56.9%) |  |
| Clear Margins  N (%) | Positive | 5 (5.2%) | 5 (9.8%) | 0.57 |
|  | Negative | 79 (82.3%) | 40 (78.4%) |  |
|  | Undetermined | 12 (12.5%) | 6 (11.8%) |  |
| Tumor size (mm),  Median (Range) | | 16.3 (4.0 - 90.0) | 21.3 (2.0 - 76.0) | 0.05 |
| Nuclear Grade  N (%) | Low | 2 (2.1%) | 2 (3.9%) | 0.31 |
|  | Moderate | 61 (63.5%) | 26 (51.0%) |  |
|  | High | 33 (34.4%) | 23 (45.1%) |  |
| Multifocality  N (%) | Present | 19 (19.8%) | 17 (33.3%) | 0.14 |
|  | Absent | 48 (50.0%) | 24 (47.1%) |  |
|  | Undetermined | 29 (30.2%) | 10 (19.6%) |  |
| Age  N (%) | <50 Years Old | 37 (38.5%) | 15 (29.4%) | 0.11 |
|  | 50-60 Years Old | 34 (35.4%) | 14 (27.5%) |  |
|  | >60 Years Old | 25 (26.1%) | 22 (43.1%) |  |
| PAM50 Subtype (Pearson Correction)  N (%) | Basal-like | 12 (12.5%) | 9 (17.6%) | 0.01 |
|  | Her2-like | 9 (9.4%) | 9 (17.6%) |  |
|  | LumA | 27 (28.1%) | 5 (9.8%) |  |
|  | LumB | 9 (9.4%) | 12 (23.5%) |  |
|  | Normal-Like | 20 (20.8%) | 5 (9.8%) |  |
|  | Undetermined | 19 (19.8%) | 11 (21.6%) |  |

**^a^**  Chi-square and t-test statistics for categorical and quantitative variables, respectively

**Table 1. Patient and tumor clinical attributes**
